# Influence of reactive balance training program characteristics on reactive balance control and fall risk: a systematic review and meta-analysis

**DOI:** 10.1101/2025.08.01.25332828

**Authors:** Hadas Nachmani, Laura K Langer, Augustine J Devasahayam, Avril Mansfield

**Author notes:** **Corresponding author:** Avril Mansfield; KITE-Toronto Rehabilitation Institute, University Health Network. Statement of ethics: Not applicable. **Author contributions:** AM conceived the concept for this review. Authors HN and AM equally contributed to the literature searches, article selection, and drafting of the manuscript. LL was responsible for the statistical analysis and data interpretation. All authors actively participated in writing the original drafts, as well as reviewing and editing the final manuscript. All authors have read and approved the published version of the manuscript. **Data availability statement:** All data are included in the manuscript.

## Abstract

**Introduction**: Diverse Reactive Balance Training (RBT) programs have been developed to address age-related deterioration in reactive balance control and increased fall risk. Despite the demonstrated effectiveness of those programs, there is significant variability in intervention characteristics (e.g., type of perturbations, total volume and intensity of training) and in study findings. It is likely that intervention effectiveness depends on features of the intervention; however, little is known about the optimal way to deliver RBT. The purpose of this systematic review and meta-analysis is to determine the optimal intervention characteristics for RBT for improving reactive balance control and preventing falls in daily life. **Methods**: We searched MEDLINE ALL (July 2023), Embase (July 2023), Physiotherapy Evidence Database (August 2023) and Cochrane (July 2023) for randomized controlled trials of RBT that reported on a measures of reactive balance control and/or falls in daily life. Results were screened by two reviewers independently to determine eligibility. The following details were extracted: study population; intervention characteristics (number of sessions in total, duration, and frequency of sessions; type, intensity and number of perturbations; description of the control intervention; and program duration), number of participants in each group; reactive balance outcomes pre- and post-intervention, and number of falls in daily life post-intervention. Risk of bias (RoB) and certainty of evidence (GRADE) were assessed. Meta-regressions were performed to explore the influence of different study components on reactive balance control and falls in daily life. **Results:** After screening 7,677 records, 32 studies were included; 25 reported a reactive balance outcome, and 19 reported falls in daily life. RoB of reactive balance control revealed main concerns arising from selection of reported results (20/25). RoB of falls in daily life had high or some concerns in the measurements of the outcome (12/19) and selection of reported results (15/19). RBT programs that included manual perturbations were associated with reduced fall rates compared to the reference (waist pull perturbations; relative risk: 0.45; 95% confidence interval: [0.22, 0.91], p=0.042). There were no other significant relationships between any other training parameters and falls in daily life or reactive balance control. Quality of evidence (GRADE) was low for both reactive balance control and falls in daily life. **Discussion:** While there was some evidence for superiority of manual perturbations over other perturbation types for fall prevention, we were unable make any definitive conclusions regarding optimal training RBT characteristics. High variability in training protocols between studies and under-reporting of intervention characteristics prevented us from making a meaningful analysis of the existing studies. Future RBT studies should provide more detailed descriptions of training protocols and include head-to-head comparisons of different training parameters (e.g., perturbation types or intensities). RBT studies should also include outcomes for both reactive balance control and falls in daily life.

## INTRODUCTION

Falls among older adults contribute to morbidity and mortality, with significant economic costs. Falls are a major contributor to trauma-related hospitalizations, and only about half of those admitted to hospital after a fall will be alive a year later [1,2]. Balance recovery abilities resulting from unexpected loss of balance decline with age and, as a result, there is an increased risk of falling in older adults [3]. Diverse fall prevention exercise programs have been developed to address this issue. In a meta-analysis, Sherrington et al. [4] found that older adults who participated in balance training programs reported 23% fewer falls and were 15% less likely to fall than those who completed a control intervention that did not include balance training. Reactive balance training (RBT) is a type of balance training that is focused on improving control of rapid balance reactions that are necessary to avoid falling after a loss of balance [5]. It is possible that this balance-recovery specific type of balance training may be even more effective for fall prevention. Indeed, another systematic review [6] found that people who participated in RBT reported 40% fewer falls than those who participated in a control intervention.

Despite the demonstrated effectiveness of RBT for preventing falls in daily life, there is significant variability in intervention characteristics (e.g., type of perturbations, total volume of training) and study findings [6]. It is likely that intervention effectiveness depends on features of the intervention; however, little is known about the optimal way to deliver RBT. The purpose of this systematic review and meta-analysis is to determine the optimal intervention characteristics for RBT (e.g., volume of training, perturbation type, perturbation intensity) for improving reactive balance control and preventing falls in daily life.

## METHODS

### Study design

This study is a systematic review and meta-analysis, based on a previous review by Devasahayam et al [6], conducted according to Cochrane guidelines [7] and reported according to the PRISMA statement and checklist [8].

### Eligibility criteria

Studies were included in this review if they met the following criteria: (1) written in English; (2) included a RBT intervention; (3) included a population at increased risk of falls (older adults; people with neurological conditions; orthopedic conditions; any other condition that increases the risk of falling); (4) included a control group that did not practice RBT; (5) random allocation of participants to RBT or control groups; and (6) outcomes included reactive balance control and/or falls in daily life after the intervention. We define RBT as a training method with a goal of improving reactive balance control; with RBT, participants may experience internal and/or external balance perturbations that cause loss of balance and evoke balance reactions (e.g., step or grasp responses).

### Information sources and search strategy

The following search engines were used for data collection-MEDLINE ALL (in Ovid, including Epub Ahead of Print, In-Process & Other Non-Indexed Citations, Ovid MEDLINE Daily; 1946 to 25 July 2023), Embase (in Ovid, including Embase Classic; 1947 to 25 July 2023), Physiotherapy Evidence Database (PEDro; searched on 1 August 2023), and Cochrane 2014 to 25 July 2023). The search strategy was developed and implemented by an information specialist. The full search strategy is included in the Appendix.

### Selection process

The titles were screened by two reviewers independently and duplicate and ineligible titles were removed (see Figure 1). The remaining abstracts were screened to determine eligibility, if eligibility was unclear from the abstract, a full text of the article was read. Disagreements regarding inclusion or exclusion were discussed and resolved. Authors of publications that met all but the last inclusion criterion (reported reactive balance measurements or fall data), were contacted to determine if these data existed. We used Endnote (X5.0.1, Thomson Reuters, Toronto, Canada) for bibliographic reference management.

**Figure 1:**
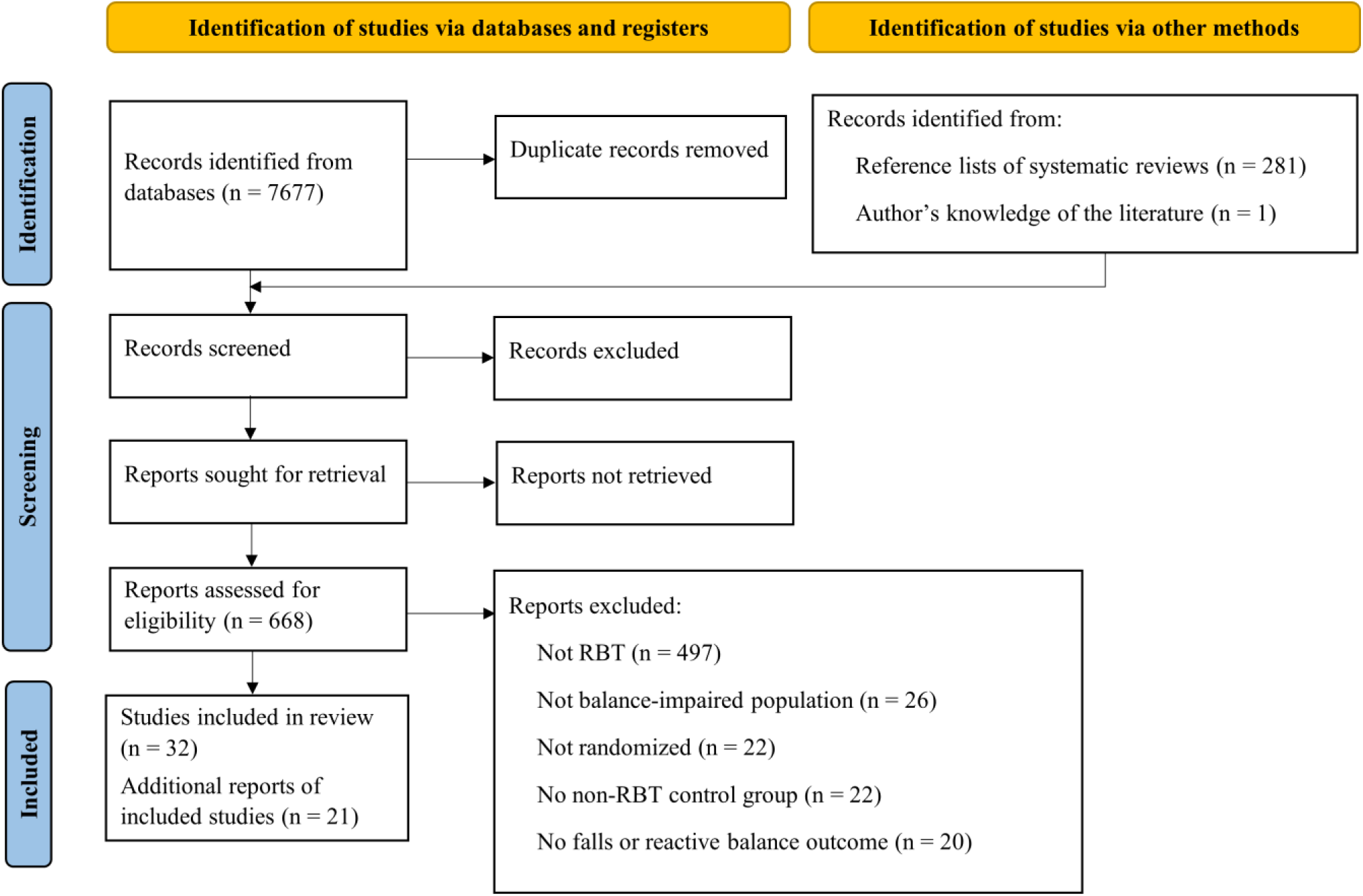
PRISMA flow diagram.

### Data collection process and data items

The following details were extracted: population of the study; RBT and control interventions (number, duration, and frequency of the sessions; type, intensity, and number of perturbations; or description of the control intervention; and program duration), number of participants in each group; reactive balance outcomes pre- and post-intervention, and data on falls in daily life post intervention. If additional information was needed, it was obtained through secondary publications or the corresponding authors. Data were compiled into a Microsoft Excel® spreadsheet. Due to variability in perturbation methods, intensity of training was not consistently defined across studies [6]. Therefore, we classified relative perturbation intensity as follows:

- Low-the goal was for participants to recover balance without stepping, or participants did not need to take a reactive step more than half of the time;
- Moderate-the goal was for participants to recover balance with just one reactive step, or participants took a single reactive step more than half of the time;
- High-the goal was for participants to recover balance with more than one reactive step, or participants took more than one reactive step more than half of the time; or
- Very high-the goal was for participants to fall, or participants fell more than half of the time.

Additionally, perturbation intensity was ‘Absolute’ if all participants experienced the same perturbation magnitude, and ‘Subjective’ as participants perceived difficulty of the perturbations was used to determine the intensity.

### Study and reporting risk of bias assessment and certainty assessment

A Risk of bias evaluation was performed by two reviewers using the revised Cochrane Risk of Bias (RoB2) Tool [9]. The assessment has five domains, risk of bias: (1) arising from the randomization process; (2) due to deviations from the intended interventions; (3) due to missing outcome data; (4) in measurement of the outcome; and (5) in selection of the reported results [9]. The assessment rated the studies as low risk/high risk/some concerns. Any disagreement between assessors was discussed and resolved. Reporting risk of bias results was through a traffic light plot of the randomized control trials; the plot illustrates the risk of bias across all five domains along with the overall risk of bias assessment for each study.

An assessment of certainty of evidence [10,11] was performed considering five domains of limitations (risk of bias, inconsistency, indirectness, imprecision, other considerations). Studies were graded by two reviewers, and limitations were labeled “not serious”/“serious”/“very serious” for each domain. The overall assessment (high, moderate, low and very low certainty of evidence) was determined based on the confidence in the estimated effects.

### Synthesis methods

To explore the influence of different study components on results of the meta-analyses, exploratory meta-regressions were performed using a restricted maximum likelihood estimation mixed effects random intercepts model for standard mean difference (SMD) meta-analysis and a minimum variance quadratic unbiased estimation mixed model with random effects for log rate ratio meta-analysis. Multivariable models were fitted using backwards variable selection methods however these models failed to achieve correctly inverted Hessian matrix and/or G-matrix was not definite positive and uni-variable meta-regressions were then performed for each identified study component. Multicolinearity was assessed for the multivariable models and log-converted number of total perps was retained instead of number of sessions. SAS (Version 9.4, SAS Institute, North Carolina, USA) for Windows was used.

## RESULTS

### Study selection

A total of 7,677 records were identified from database searching. After removing duplicates (n=2,878), 5,085 records were screened for eligibility. Of these, 53 reports of 32 studies were included in this review (Figure 1). Appendix B includes details for all reports.

### Study characteristics

The characteristics of the studies are presented in Table 1. Samples included in this review were: healthy older adults [12–24]; older adults with mixed diagnoses [25–29]; people with Parkinson’s disease [30–36]; people with chronic [37–39] and sub-acute [40] stroke; people with multiple sclerosis [41,42]; and people with incomplete spinal cord injury [43]. Twenty studies used surface translation perturbations, 5 used manual perturbations, 5 used overground perturbations (slip and trip walkway), and 2 studies used waist pull perturbations. Perturbation directions varied from a single direction (i.e., backward (n=4) or forward (n=1) only), a single axis (antero-posterior (n=13) or mediolateral (n=1)), and multidirectional (n=13). Perturbations were delivered during walking in 13 studies, standing in 5 studies and various posture in 9 studies. Perturbation intensities were low (n=1), moderate (n=11), high (n=5), absolute (n=7), and subjective (n=4). Total training duration ranged from 15 to 1800 minutes (mean: 573 minutes, standard deviation: 484 minutes), and the total number of perturbations was 16 to 1440 (mean: 539, standard deviation: 493). Table 2 presents the individual results of reactive balance control outcomes from each study. Table 3 summarizes the rate ratios of falls in daily life as reported in the studies.

**Table 1:** Study characteristics.

| Study | Country | Sample | Reactive balance training details<br>Type | Direction | Posture | Intensity | Duration-min<br>(program) | Number of<br>perturbations | Type of control<br>group | Reactive<br>balance<br>control<br>outcome | Fall<br>rate |
| --- | --- | --- | --- | --- | --- | --- | --- | --- | --- | --- | --- |
| Shimada<br>2004 | Japan | Older<br>adults,<br>mixed<br>diagnoses | Surface<br>translation | Backward | Walking | Absolute | 600 | . | Usual care | - | Yes |
| Protas<br>2005 | USA | Parkinson's<br>disease | Surface<br>translation | Multi-directional | Standing | High | 1440 | 720 | No intervention | - | Yes |
| Marigold<br>2005 | Canada | Chronic<br>stroke | Manual | Multi-directional | Standing | Moderate | 1800 | . | Stretching/weight-<br>shifting exercises | In-lab falls | Yes |
| Bieryla<br>2007 | USA | Healthy<br>older adults | Overground | Forward | Walking | Moderate | 15 | 20 | Treadmill walking | Maximum trunk<br>angle following<br>trip during<br>walking | No |
| Maki<br>2008 | Canada | Older<br>adults,<br>mixed<br>diagnoses | Surface<br>translation | Multi-directional | Various | Moderate | 540 | 864 | Voluntary<br>stepping and<br>grasping<br>movements | - | Yes |
| Smania<br>2010 | Italy | Parkinson's<br>disease | Manual | Multi-directional | Various | Low | 1050 | . | Joint mobilization,<br>muscle<br>stretching, and<br>motor<br>coordination<br>exercises | UPDRS total<br>score | Yes |
| Mansfield<br>2010 | Canada | Healthy<br>older adults | Surface<br>translation | Multi-directional | Various | Moderate | 540 | 1008 | Flexibility<br>exercises and<br>guided meditation | Frequency of<br>multi-step<br>reactions | Yes |
| Parijat<br>2012 | USA | Healthy<br>older adults | Overground | Backward | Walking | Absolute | 37.5 | 24 | Unperturbed<br>walking | In-lab falls | No |
| Shen<br>2012 | China | Parkinson's<br>disease | Surface<br>translation | Multi-directional | Standing | Absolute | 720 | 480 | Resistance<br>training | UPDRS-PG<br>(posture and<br>gait sub-score) | No |
| Lurie<br>2013 | USA | Older<br>adults,<br>mixed<br>diagnoses | Surface<br>translation | Antero-posterior | Walking | . | . | . | Usual care<br>(balance and gait<br>training) | - | Yes |

| Study | Country | Sample | Reactive balance training details |  |  |  | Duration-min<br>(program) | Number of perturbations | Type of control group | Reactive balance control outcome | Fall rate |
| --- | --- | --- | --- | --- | --- | --- | --- | --- | --- | --- | --- |
|  |  |  | Type | Direction | Posture | Intensity |  |  |  |  |  |
| Pai 2014 | USA | Older adults, mixed diagnoses | Overground | Backward | Walking | Absolute | . | 24 | No intervention | - | Yes |
| Schlenstedt 2015 | Germany | Parkinson's disease | Manual | Multi-directional | Various | . | 840 | . | Resistance training | Fullerton Advanced Balance scale | No |
| Shen 2015 | China | Parkinson's disease | Surface translation | Antero-posterior | Walking | Moderate | 1440 | . | Resistance training | Latency of balance reactions | Yes |
| Kurz 2016 | Israel | Healthy older adults | Surface translation | Multi-directional | Walking | Moderate | 480 | 1100 | Treadmill walking | Performance-Oriented Mobility Assessment - balance sub score | Yes |
| Steib 2017 | Germany | Parkinson's disease | Surface translation | Antero-posterior | Walking | . | 640 | . | Treadmill walking | Mini-BEST reactive sub-score | No |
| Mansfield 2018 | Canada | Chronic stroke | Manual | Multi-directional | Various | High | 720 | 720 | Conventional balance training | Mini-BEST reactive sub-score | Yes |
| Aviles 2019 | USA | Healthy older adults | Surface translation | Antero-posterior | Standing | High | 360 | 480 | Tai Chi | Custom reactive balance rating | Yes |
| Handelzalts 2019 | Israel | Sub-acute stroke | Surface translation | Antero-posterior | Various | High | 360 | 432 | Weight shifting and gait training | Fall threshold (in-lab) | No |
| Okubo 2019 | Australia | Healthy older adults | Overground | Antero-posterior | Walking | Subjective | 120 | 60 | Overground walking | In-lab falls | No |
| Esmaeili 2020 | Canada | Chronic stroke | Surface translation | Antero-posterior | Walking | Moderate | . | 234 | Treadmill walking | Mini-BEST | No |
| Lurie 2020 | USA | Older adults, mixed diagnoses | Surface translation | Antero-posterior | Walking | . | 562.5 | . | Usual care (balance and gait training) | - | Yes |
|  |  |  | Type | Direction | Posture | Intensity |  |  |  |  |  |
| Rieger 2020 | Netherlands | Healthy older adults | Surface translation | Antero-posterior | Walking | Absolute | . | 16 | Treadmill walking | Deviation in trunk velocity from unperturbed walking | No |
| Rogers 2021 | USA | Healthy older adults | Waist pull | Medio-lateral | Standing | Moderate | . | 1440 | Hip strengthening | Multi-step threshold | Yes |
| Unger 2021 | Canada | Incomplete spinal cord injury | Manual | Multi-directional | Various | Moderate | 1440 | 1440 | 'Conventional' intense balance training | Behavioural response score following perturbations | Yes |
| Monjezi 2022 | Iran | Multiple sclerosis | Waist pull | Backward | Various | High | 240 | 600 | Standing balance exercises and Treadmill walking | Latency of postural responses | Yes |
| Wang 2022 | USA | Healthy older adults | Surface translation | Antero-posterior | Walking | Absolute | 30 | 40 | Treadmill walking | - | Yes |
| Brull 2023 | Germany | Healthy older adults | Surface translation | Multi-directional | Walking | Moderate | 432 | 1296 | No intervention | Stepping Threshold Test | No |
| de Souza 2023 | Brazil | Parkinson's disease | Surface translation | Multi-directional | Standing | Absolute | 440 | 1024 | Resistance training | In-lab near falls | No |
| Gerards 2023 | Netherlands | Healthy older adults | Surface translation | Multi-directional | Various | Subjective | 90 | . | Usual care | Mini-BEST reactive sub-score | No |
| Nørgaard 2023 | Denmark | Healthy older adults | Surface translation | Antero-posterior | Walking | Subjective | 40 | 80 | Treadmill walking | In-lab falls | Yes |
| Okubo 2023 | Australia | Multiple Sclerosis | Overground | Antero-posterior | Walking | Subjective | 100 | 48 | Sham obstacle avoidance training | In-lab falls | No |
| Rieger 2023 | Netherlands | Healthy older adults | Surface translation | Antero-posterior | Walking | Moderate | 240 | 256 | Treadmill walking with cognitive task | Mini-BEST reactive sub-score | Yes |

BEST: Balance Evaluation Systems Test; UPDRS: Unified Parkinson's Disease rating scale

**Table 2:**
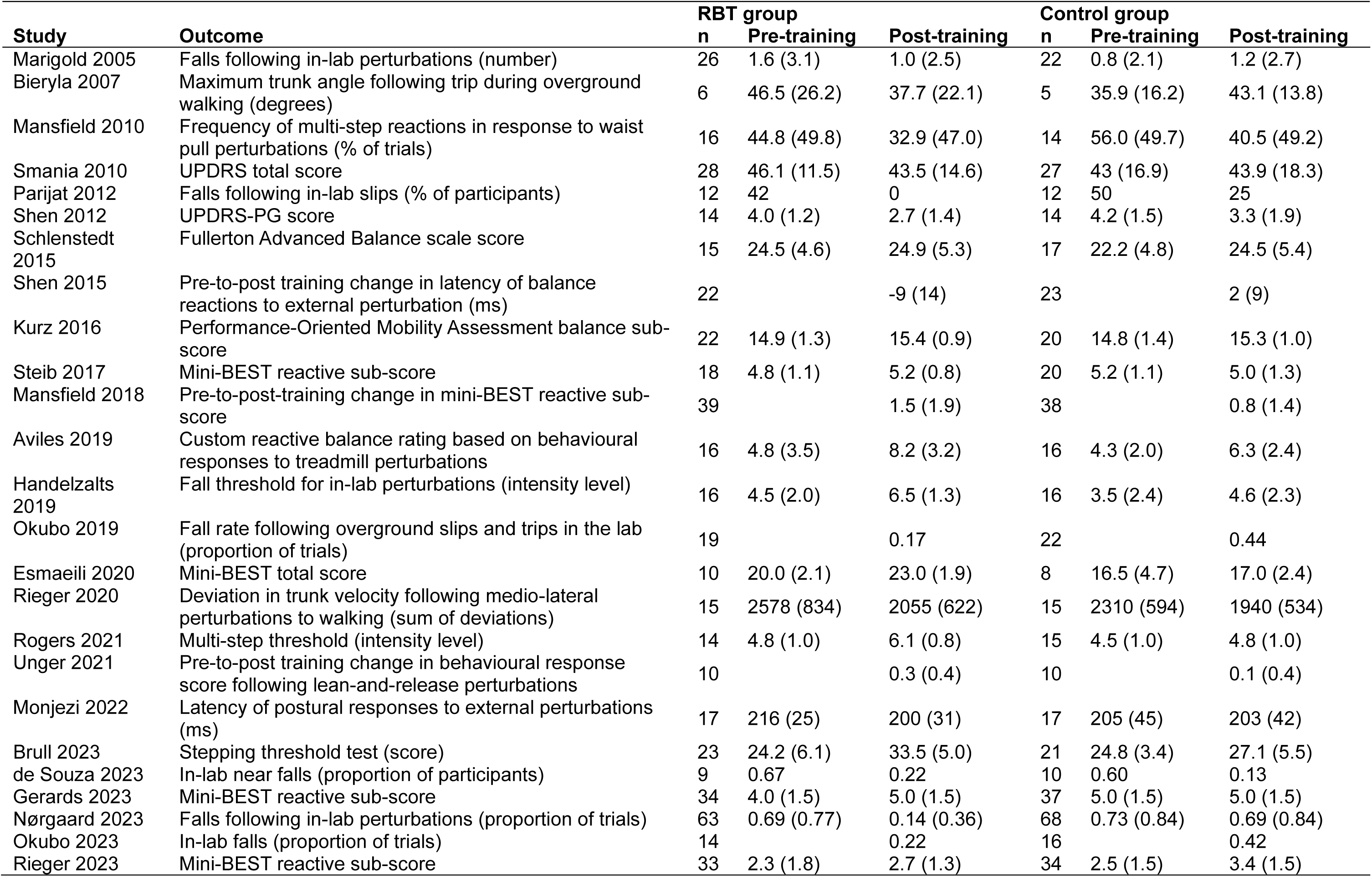
Results of studies reporting on reactive balance control outcomes. Values presented are means with standard deviations in parentheses.

| Study | Outcome | RBT group |  |  | Control group |  |  |
| --- | --- | --- | --- | --- | --- | --- | --- |
|  |  | n | Pre-training | Post-training | n | Pre-training | Post-training |
| Marigold 2005 | Falls following in-lab perturbations (number) | 26 | 1.6 (3.1) | 1.0 (2.5) | 22 | 0.8 (2.1) | 1.2 (2.7) |
| Bieryla 2007 | Maximum trunk angle following trip during overground walking (degrees) | 6 | 46.5 (26.2) | 37.7 (22.1) | 5 | 35.9 (16.2) | 43.1 (13.8) |
| Mansfield 2010 | Frequency of multi-step reactions in response to waist pull perturbations (% of trials) | 16 | 44.8 (49.8) | 32.9 (47.0) | 14 | 56.0 (49.7) | 40.5 (49.2) |
| Smania 2010 | UPDRS total score | 28 | 46.1 (11.5) | 43.5 (14.6) | 27 | 43 (16.9) | 43.9 (18.3) |
| Parijat 2012 | Falls following in-lab slips (% of participants) | 12 | 42 | 0 | 12 | 50 | 25 |
| Shen 2012 | UPDRS-PG score | 14 | 4.0 (1.2) | 2.7 (1.4) | 14 | 4.2 (1.5) | 3.3 (1.9) |
| Schlenstedt 2015 | Fullerton Advanced Balance scale score | 15 | 24.5 (4.6) | 24.9 (5.3) | 17 | 22.2 (4.8) | 24.5 (5.4) |
| Shen 2015 | Pre-to-post training change in latency of balance reactions to external perturbation (ms) | 22 |  | -9 (14) | 23 |  | 2 (9) |
| Kurz 2016 | Performance-Oriented Mobility Assessment balance sub-score | 22 | 14.9 (1.3) | 15.4 (0.9) | 20 | 14.8 (1.4) | 15.3 (1.0) |
| Steib 2017 | Mini-BEST reactive sub-score | 18 | 4.8 (1.1) | 5.2 (0.8) | 20 | 5.2 (1.1) | 5.0 (1.3) |
| Mansfield 2018 | Pre-to-post-training change in mini-BEST reactive sub-score | 39 |  | 1.5 (1.9) | 38 |  | 0.8 (1.4) |
| Aviles 2019 | Custom reactive balance rating based on behavioural responses to treadmill perturbations | 16 | 4.8 (3.5) | 8.2 (3.2) | 16 | 4.3 (2.0) | 6.3 (2.4) |
| Handelzalts 2019 | Fall threshold for in-lab perturbations (intensity level) | 16 | 4.5 (2.0) | 6.5 (1.3) | 16 | 3.5 (2.4) | 4.6 (2.3) |
| Okubo 2019 | Fall rate following overground slips and trips in the lab (proportion of trials) | 19 |  | 0.17 | 22 |  | 0.44 |
| Esmaeili 2020 | Mini-BEST total score | 10 | 20.0 (2.1) | 23.0 (1.9) | 8 | 16.5 (4.7) | 17.0 (2.4) |
| Rieger 2020 | Deviation in trunk velocity following medio-lateral perturbations to walking (sum of deviations) | 15 | 2578 (834) | 2055 (622) | 15 | 2310 (594) | 1940 (534) |
| Rogers 2021 | Multi-step threshold (intensity level) | 14 | 4.8 (1.0) | 6.1 (0.8) | 15 | 4.5 (1.0) | 4.8 (1.0) |
| Unger 2021 | Pre-to-post training change in behavioural response score following lean-and-release perturbations | 10 |  | 0.3 (0.4) | 10 |  | 0.1 (0.4) |
| Monjezi 2022 | Latency of postural responses to external perturbations (ms) | 17 | 216 (25) | 200 (31) | 17 | 205 (45) | 203 (42) |
| Brull 2023 | Stepping threshold test (score) | 23 | 24.2 (6.1) | 33.5 (5.0) | 21 | 24.8 (3.4) | 27.1 (5.5) |
| de Souza 2023 | In-lab near falls (proportion of participants) | 9 | 0.67 | 0.22 | 10 | 0.60 | 0.13 |
| Gerards 2023 | Mini-BEST reactive sub-score | 34 | 4.0 (1.5) | 5.0 (1.5) | 37 | 5.0 (1.5) | 5.0 (1.5) |
| Nørgaard 2023 | Falls following in-lab perturbations (proportion of trials) | 63 | 0.69 (0.77) | 0.14 (0.36) | 68 | 0.73 (0.84) | 0.69 (0.84) |
| Okubo 2023 | In-lab falls (proportion of trials) | 14 |  | 0.22 | 16 |  | 0.42 |
| Rieger 2023 | Mini-BEST reactive sub-score | 33 | 2.3 (1.8) | 2.7 (1.3) | 34 | 2.5 (1.5) | 3.4 (1.5) |

**Table 3:** Falls in daily life.

| Study | RBT group |  | Control group |  | Rate ratio |
| --- | --- | --- | --- | --- | --- |
|  | n | Number of falls | n | Number of falls |  |
| Shimada 2004 | 15 | 8 | 11 | 11 | 0.533 |
| Marigold 2005 | 19 | 25 | 21 | 75 | 0.368 |
| Protas 2005 | 9 | 10 | 9 | 23 | 0.435 |
| Maki 2008 | 4 | 4 | 4 | 4 | 1.000 |
| Mansfield 2010 | 16 | 6 | 15 | 4 | 1.406 |
| Smania 2010 | 28 | 36 | 27 | 111 | 0.313 |
| Lurie 2013 | 26 | 10 | 33 | 32 | 0.397 |
| Pai 2014 | 102 | 17 | 96 | 32 | 0.500 |
| Shen 2015 | 22 | 10 | 23 | 18 | 0.581 |
| Kurz 2016 | 21 | 12 | 19 | 18 | 0.603 |
| Mansfield 2018 | 41 | 53 | 42 | 64 | 0.848 |
| Aviles 2019 | 16 | 4 | 16 | 5 | 0.800 |
| Lurie 2020 | 218 | 686 | 212 | 603 | 1.106 |
| Rogers 2021 | 18 | 7 | 15 | 4 | 1.444 |
| Unger 2021 | 10 | 10 | 10 | 21 | 0.476 |
| Monjezi 2022 | 17 | 46 | 17 | 64 | 0.719 |
| Wang 2022 | 70 | 37 | 63 | 28 | 1.189 |
| Nørgaard 2023 | 63 | 62 | 68 | 78 | 0.780 |
| Rieger 2023 | 33 | 10 | 34 | 13 | 0.793 |

### Risk of bias

Figure 2 presents the risk of bias for studies reporting reactive balance outcomes (n = 25). Six studies were rated as high risk, 14 had some concerns, and five were assessed as low risk of bias. The main concerns arose from the selection of reported results (20 of 25) and deviations from the intended interventions (8 of 25). In contrast, most studies showed low risk of bias in randomization (19 of 25), missing data (24 of 25), and outcome measurement (21 of 25).

**Figure 2:**
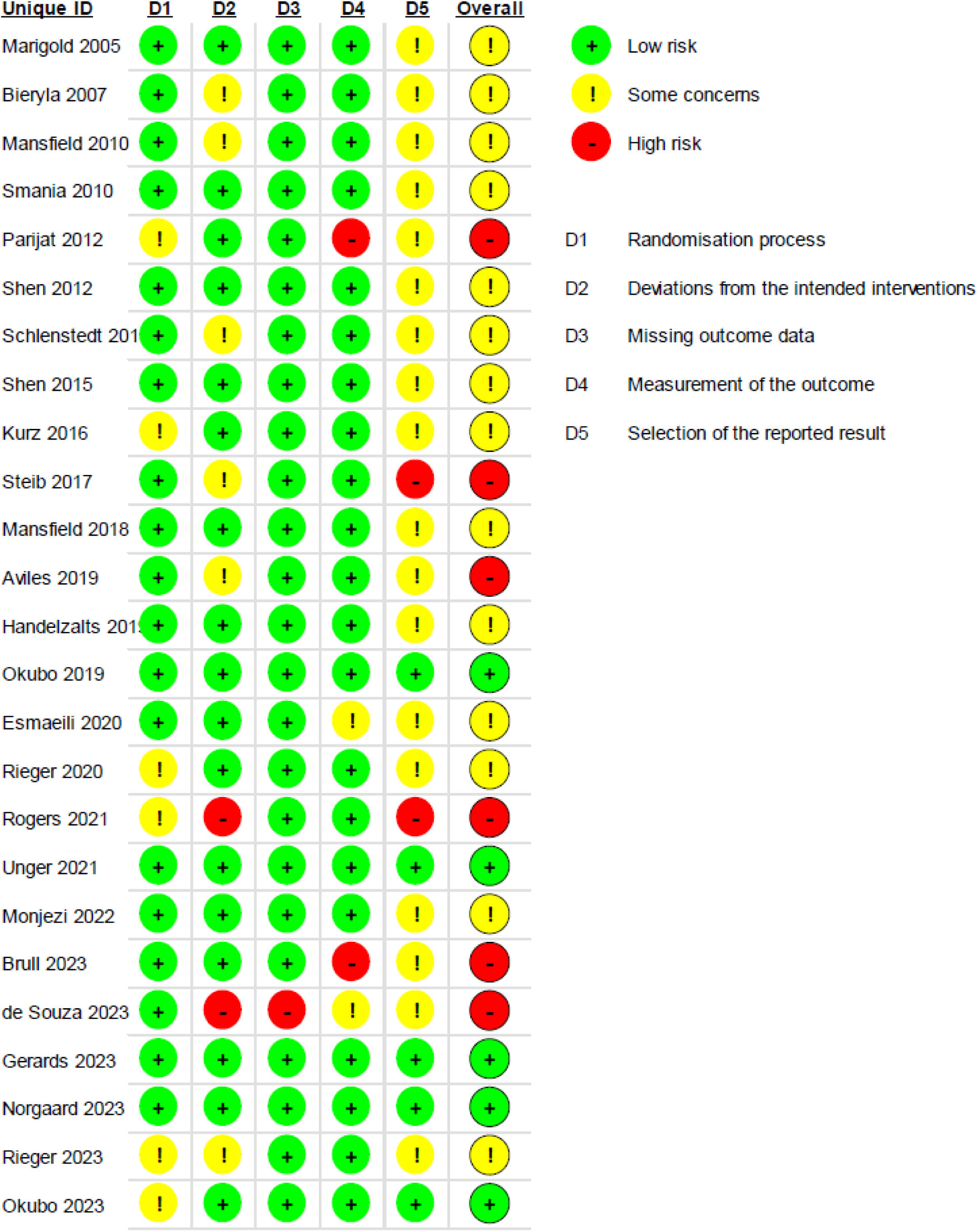
Risk of bias for studies reporting a reactive balance control outcome.

Figure 3 presents the risk of bias for the 19 studies reporting falls in daily life. Nine studies were rated as high risk, eight had some concerns, and only two were assessed as low risk. These findings primarily reflect high or some concerns in the randomization process (7 of 19 studies), outcome measurement (12 of 19), and selection of reported results (15 of 19). In contrast, most studies showed low risk of bias for missing data (18 of 19) and deviations from intended interventions (15 of 19).

**Figure 3:**
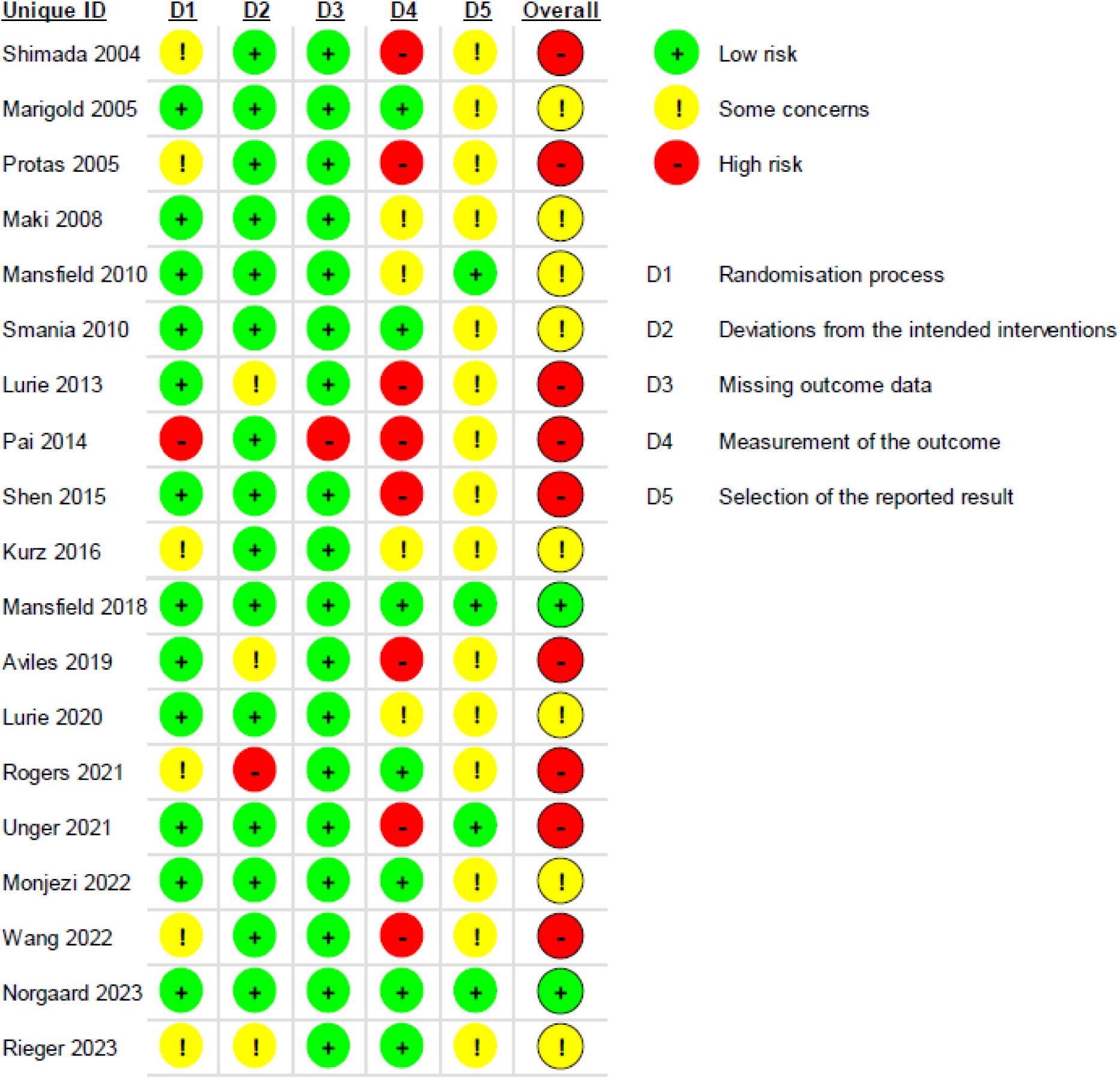
Risk of bias for studies reporting falls in daily life.

### Meta regression

Table 4 describes the meta regression analysis of the influence of each training parameter on reactive balance outcomes and falls in daily life. RBT programs that included manual perturbations were associated with reduced falls in daily life compared to the reference (waist pull perturbations; relative risk: 0.45; 95% confidence interval: [0.22, 0.91], p=0.042). There were no other significant relationships between any other training parameters and falls in daily life or reactive balance control outcomes.

**Table 4:**
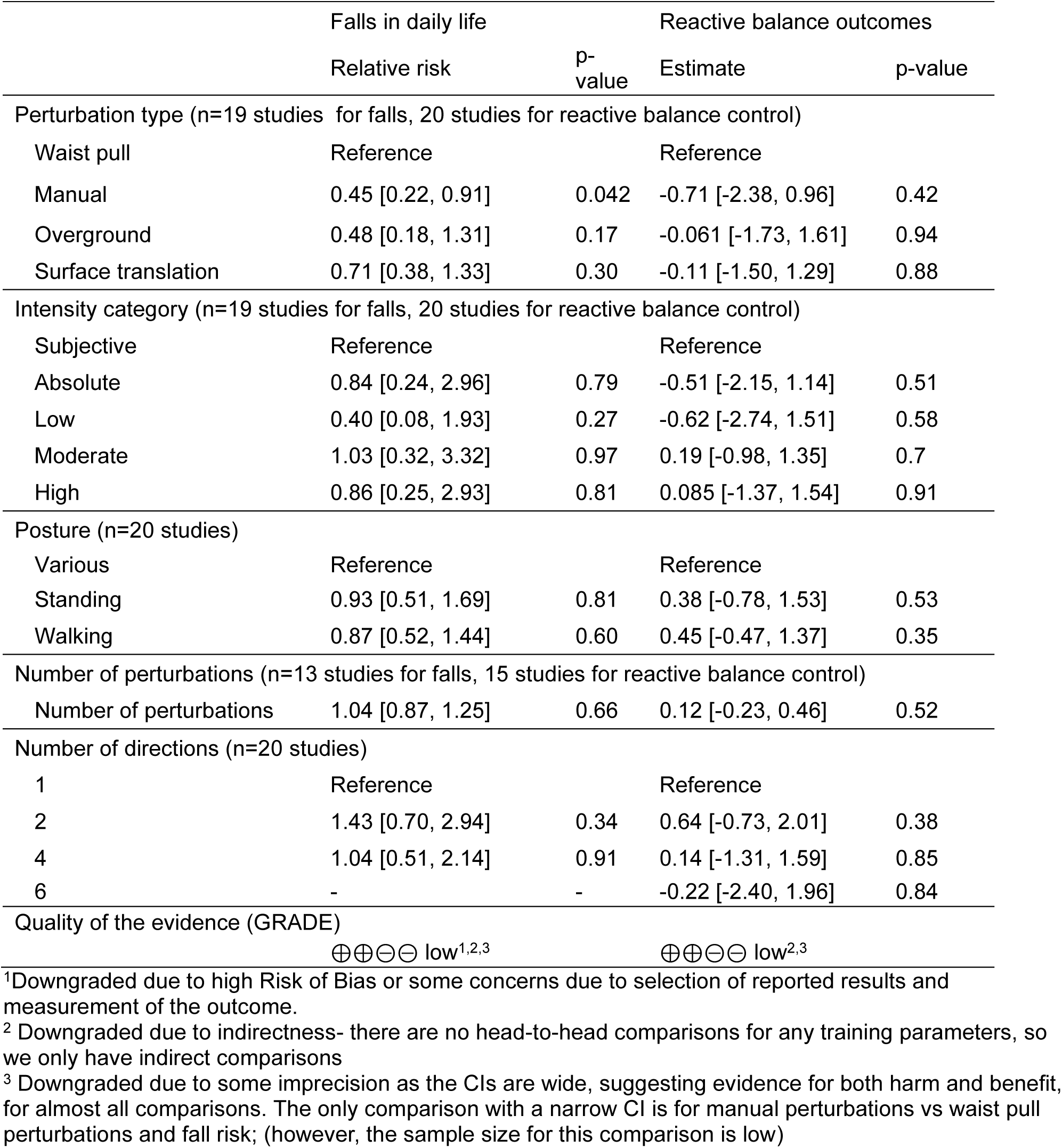
Univariable meta-regressions for intervention components. Values presented are estimates (relative risk for falls in daily life, slope for reactive balance outcomes) with confidence intervals in brackets and associated p-values.

### GRADE quality of evidence

The evidence quality for reactive balance outcome and fall in daily life outcome were rated as low according to the GRADE approach. The studies included in this meta-analysis are RCTs, which provide high-quality evidence. However, there were some limitations that downgraded the quality of the evidence (Table 4).

## DISCUSSION

RBT has been widely shown to improve reactive balance control [44] and reduce falls in daily life [6]. This review aimed to determine the optimal RBT intervention characteristics for improving reactive balance control and preventing falls in daily life. However, we were unable to identify key training program characteristics that contribute to improved training effects.

Our analysis suggests that RBT using manual perturbations may be superior to waist-pull perturbations for preventing falls in daily life. Among people with significant balance impairments, such as those with chronic stroke [45] or long-term care residents [46], a large number of falls occur due to “incorrect weight transfer” (i.e., loss of balance during voluntary movement), rather than due to external forces or environmental hazards (e.g., slips or trips). Manual RBT includes both external and internal perturbations (i.e., “incorrect weight transfers”); if this training mimics how people typically lose balance prior to a fall in daily life, this may explain the apparent benefits of manual RBT for preventing falls. However, it is possible that other differences between studies account for the apparent significant reduction in fall rates for manual RBT. For example, all manual RBT studies included people with neurological conditions (stroke, Parkinson’s disease, and spinal cord injury); it is possible that large training effects for falls in daily life were observed for these studies due to the high baseline fall rate among these groups.

No studies in our review compared different RBT intensities, and few studies [12,16,17,20,21,24,27–29,31,40] reported the intensity of training. Two previous studies that included healthy young adults found that higher intensity RBT led to greater improvements in reactive balance control than lower intensity training [47,48]. Conversely, a more recent study comparing different RBT intensity schedules among healthy young adults found no difference in training effect between the low-to-high intensity training schedule and the group that always completed RBT at a high intensity [49]. For other types of exercise and components of fitness, shorter high intensity training sessions are often found to lead to similar or improved outcomes compared to longer sessions of moderate intensity training [50]. Future studies should compare RBT of different intensities to determine if the same is true for RBT. In our analysis, we expected that a higher training volume (i.e., more perturbations) would lead to better outcomes. This expectation is based on previous studies showing that more practice generally leads to better outcomes [51]. However, some RBT studies have demonstrated significant effects with 1-3 relatively short training sessions [19,52]. While these studies used absolute (all participants experienced the same intensity) and subjective RBT intensities, the intensity of training in these studies may have been high to very high (i.e., evident by reports of falls into the safety harness in these studies). Therefore, high intensity training may explain the significant training effects with a short duration of training. This hypothesis will need to be tested in future studies.

We anticipated that a more variable practice, incorporating a variety of postures, directions, and fall-risk situations, would be more effective for improving reactive balance control and reducing fall rates [53]. However, we did not observe greater training effects for programs that included greater variability in postures or directions. Walking is one of the most frequent activities at the time of a fall [38,46]. However, in our study, training programs that focused on walking were not more effective than programs that provided perturbations to stance only, or that included various postures.

Our findings are similar to those of Farlie et al. [54], who conducted a meta-analysis of general balance training programs, and found no association between the components of the ‘FITT’ principle (frequency, intensity, type, and time) and improved balance outcomes following balance training. Similar to this previous analysis, we also found that no evidence that specific RBT program characteristics led to superior training effects. Farlie et al. [54] speculated that their findings may be partially explained by incomplete reporting of training characteristics in published studies, which may also be the case for our review. Lack of reporting key study characteristics also reduced the available sample size for our analyses [54]. Less than half of the studies (14/32) reported all training characteristics in a way that could be meaningfully incorporated into meta-analysis. Due to the small sample size and high variability in study designs (e.g., in participant characteristics, control group interventions), we were unable to conduct multi-variable meta-regression to examine the effect of multiple training characteristics in combination with each other. The Template for Intervention Description and Replication (TIDieR) [55] and Consensus on Exercise Reporting Template (CERT; [56]) are guidelines for reporting intervention characteristics in clinical trials. Theses reporting guidelines should be used by authors of RBT studies to ensure that intervention characteristics are reported fully, and use of these guidelines should be enforced by journal reviewers and editors. More consistent use of these reporting guidelines will not only facilitate meta-analyses, like ours, but also establishing clinical practice guidelines and facilitating translation of effective interventions into clinical practice.

This review included a wide range of study designs, with different types of training methods, substantial variability in training parameters, and diverse outcome measures; however, despite its comprehensive scope, definitive conclusions could not be drawn.

Despite including only RCTs, the quality of evidence for reactive balance and falls in daily life was low, due to wide confidence intervals, lack of direct comparisons, and risk of bias—posing an additional limitation, as the true effect may differ from our findings.

We suggest that future studies develop training programs based on existing training protocols, with appropriate modifications. Given the wide variety of training programs available, it is important to design studies that rely on existing research to validate their effects. There is a need to conduct an in-depth investigation into the guiding principles for designing RBT, i.e., for example, identifying the most effective perturbation parameters for fall prevention or determining the optimal intervention duration for improving balance reactions. Future studies should provide more detailed reports of the training protocols.

## Data Availability

All data are included in the manuscript.

## APPENDIX 1: Search strategies

### Ovid MEDLINE(R) ALL <1946 to July 25, 2023>

**1** (perturb* adj3 (train* or rehab* or exercis*)).tw,kf. (485)
**2** (platform* adj2 (train* or rehab* or exercis*)).tw,kf. (881)
**3** (surface translation? adj2 (train* or rehab* or exercis*)).tw,kf. (1)
**4** (“dynamic balanc*” adj4 (train* or rehab* or exercis*)).tw,kf. (281)
**5** (“dynamic stabil*” adj4 (train* or rehab* or exercis*)).tw,kf. (57)
**6** (“reactive balanc*” adj4 (train* or rehab* or exercis*)).tw,kf. (47)
**7** ((slip? or slipping) adj2 (train* or rehab* or exercis*)).tw,kf. (52)
**8** ((trip? or tripping) adj2 (train* or rehab* or exercis*)).tw,kf. (72)
**9** 1 or 2 or 3 or 4 or 5 or 6 or 7 or 8 (1793)
**10** ((step? or stepping) adj4 (train* or rehab* or exercis*)).tw,kf. (4149)
**11** (gait adj4 (train* or rehab* or exercis*)).tw,kf. (4766)
**12** ((walk? or walking) adj4 (train* or rehab* or exercis*)).tw,kf. (7213)
**13** (locomot* adj4 (train* or rehab* or exercis*)).tw,kf. (1486)
**14** (balanc* adj4 (train* or rehab* or exercis*)).tw,kf. (7629)
**15** (stabil* adj4 (train* or rehab* or exercis*)).tw,kf. (3133)
**16** (agil* adj4 (train* or rehab* or exercis*)).tw,kf. (341)
**17** 10 or 11 or 12 or 13 or 14 or 15 or 16 (25952)
**18** perturb*.tw,kf. (132693)
**19** platform*.tw,kf. (251308)
**20** surface translation?.tw,kf. (185)
**21** destabil*.tw,kf. (40535)
**22** compensat*.tw,kf. (185330)
**23** react*.tw,kf. (2140411)
**24** dynamic balanc*.tw,kf. (4668)
**25** dynamic stabil*.tw,kf. (2629)
**26** 18 or 19 or 20 or 21 or 22 or 23 or 24 or 25 (2694202)
**27** 17 and 26 (3526)
**28** 9 or 27 (4747)
**29** randomized controlled trial.pt. (597488)
**30** controlled clinical trial.pt. (95390)
**31** random*.ab. (1393741)
**32** placebo.ab. (240241)
**33** trial.ab. (658048)
**34** groups.ab. (2548683)
**35** 29 or 30 or 31 or 32 or 33 or 34 (3943449)
**36** exp animals/ not humans.sh. (5142398)
**37** 35 not 36 (3411214)
**38** 28 and 37 (2063)
**39** limit 38 to english language (2021)
**40** (“20220318” or “20220319” or 2022032* or 2022033* or 202204* or 202205* or 202206* or 202207* or 202208* or 202209* or 20221* or 2023*).dt,ez,da. (2502650)
**41** 39 and 40 (323)

### Cochrane Central Register of Controlled Trials <JUNE 2023>

1. (perturb* adj3 (train* or rehab* or exercis*)).tw,hw. (240)
2. (platform* adj2 (train* or rehab* or exercis*)).tw,hw. (276)
3. (surface translation? adj2 (train* or rehab* or exercis*)).tw,hw. (1)
4. (“dynamic balanc*” adj4 (train* or rehab* or exercis*)).tw,hw. (307)
5. (“dynamic stabil*” adj4 (train* or rehab* or exercis*)).tw,hw. (62)
6. (“reactive balanc*” adj4 (train* or rehab* or exercis*)).tw,hw. (34)
7. ((slip? or slipping) adj2 (train* or rehab* or exercis*)).tw,hw. (31)
8. ((trip? or tripping) adj2 (train* or rehab* or exercis*)).tw,hw. (21)
9. 1 or 2 or 3 or 4 or 5 or 6 or 7 or 8 (899)
10. ((step? or stepping) adj4 (train* or rehab* or exercis*)).tw,hw. (1454)
11. (gait adj4 (train* or rehab* or exercis*)).tw,hw. (3407)
12. ((walk? or walking) adj4 (train* or rehab* or exercis*)).tw,hw. (6541)
13. (locomot* adj4 (train* or rehab* or exercis*)).tw,hw. (495)
14. (balanc* adj4 (train* or rehab* or exercis*)).tw,hw. (6011)
15. (stabil* adj4 (train* or rehab* or exercis*)).tw,hw. (2550)
16. (agil* adj4 (train* or rehab* or exercis*)).tw,hw. (293)
17. 10 or 11 or 12 or 13 or 14 or 15 or 16 (17473)
18. perturb*.tw,hw. (1914)
19. platform*.tw,hw. (12113)
20. surface translation?.tw,hw. (24)
21. destabil*.tw,hw. (281)
22. compensat*.tw,hw. (9133)
23. react*.tw,hw. (168708)
24. dynamic balanc*.tw,hw. (1750)
25. dynamic stabil*.tw,hw. (311)
26. 18 or 19 or 20 or 21 or 22 or 23 or 24 or 25 (191095)
27. 17 and 26 (2703)
28. 9 or 27 (3049)
29. limit 28 to english language (3000)
30. (“202203” or “202204” or “202205” or “202206” or “202207” or “202208” or “202209” or “202210” or “202211” or “202212” or 2023*).up. (645758)
31. 29 and 30 (1188)

### Embase Classic+Embase <1947 to 2023 July 25>

1. (perturb* adj3 (train* or rehab* or exercis*)).tw,kw. (573)
2. (platform* adj2 (train* or rehab* or exercis*)).tw,kw. (1333)
3. (surface translation? adj2 (train* or rehab* or exercis*)).tw,kw. (1)
4. (“dynamic balanc*” adj4 (train* or rehab* or exercis*)).tw,kw. (404)
5. (“dynamic stabil*” adj4 (train* or rehab* or exercis*)).tw,kw. (102)
6. (“reactive balanc*” adj4 (train* or rehab* or exercis*)).tw,kw. (55)
7. ((slip? or slipping) adj2 (train* or rehab* or exercis*)).tw,kw. (60)
8. ((trip? or tripping) adj2 (train* or rehab* or exercis*)).tw,kw. (87)
9. 1 or 2 or 3 or 4 or 5 or 6 or 7 or 8 (2519)
10. ((step? or stepping) adj4 (train* or rehab* or exercis*)).tw,kw. (6088)
11. (gait adj4 (train* or rehab* or exercis*)).tw,kw. (7578)
12. ((walk? or walking) adj4 (train* or rehab* or exercis*)).tw,kw. (11634)
13. (locomot* adj4 (train* or rehab* or exercis*)).tw,kw. (2226)
14. (balanc* adj4 (train* or rehab* or exercis*)).tw,kw. (11230)
15. (stabil* adj4 (train* or rehab* or exercis*)).tw,kw. (4223)
16. (agil* adj4 (train* or rehab* or exercis*)).tw,kw. (454)
17. 10 or 11 or 12 or 13 or 14 or 15 or 16 (38938)
18. perturb*.tw,kw. (141786)
19. platform*.tw,kw. (331193)
20. surface translation?.tw,kw. (194)
21. destabil*.tw,kw. (46886)
22. compensat*.tw,kw. (240825)
23. react*.tw,kw. (2709764)
24. dynamic balanc*.tw,kw. (5895)
25. dynamic stabil*.tw,kw. (3055)
26. 18 or 19 or 20 or 21 or 22 or 23 or 24 or 25 (3403035)
27. 17 and 26 (5146)
28. 9 or 27 (6887)
29. Randomized controlled trial/ (777853)
30. Controlled clinical study/ (470945)
31. random*.ti,ab. (1967546)
32. randomization/ (98260)
33. intermethod comparison/ (298898)
34. placebo.ti,ab. (367794)
35. (compare or compared or comparison).ti. (632261)
36. ((evaluated or evaluate or evaluating or assessed or assess) and (compare or compared or comparing or comparison)).ab. (2755693)
37. (open adj label).ti,ab. (107739)
38. ((double or single or doubly or singly) adj (blind or blinded or blindly)).ti,ab. (277686)
39. double blind procedure/ (211523)
40. parallel group?.ti,ab. (31879)
41. (crossover or cross over).ti,ab. (125116)
42. ((assign* or match or matched or allocation) adj5 (alternate or group? or intervention? or patient? or subject? or participant?)).ti,ab. (413902)
43. (assigned or allocated).ti,ab. (488960)
44. (controlled adj7 (study or design or trial)).ti,ab. (449776)
45. (volunteer or volunteers).ti,ab. (287296)
46. human experiment/ (635406)
47. trial.ti. (405231)
48. or/29-47 (6344646)
49. (random* adj sampl* adj7 (“cross section*” or questionnaire? or survey? or database?)).ti,ab. not (comparative study/ or controlled study/ or randomi#ed controlled.ti,ab. or randomly assigned.ti,ab.) (9118)
50. Cross-sectional study/ not (randomized controlled trial/ or controlled clinical study/ or controlled study/ or randomi#ed controlled.ti,ab. or control group?.ti,ab.) (355810)
51. (((case adj control*) and random*) not randomi#ed controlled).ti,ab. (21396)
52. (Systematic review not (trial or study)).ti. (254679)
53. (nonrandom* not random*).ti,ab. (18961)
54. “Random field*”.ti,ab. (2945)
55. (review.ab. and review.pt.) not trial.ti. (1117119)
56. “we searched”.ab. and (review.ti. or review.pt.) (48618)
57. “update review”.ab. (135)
58. (databases adj4 searched).ab. (61279)
59. (rat or rats or mouse or mice or swine or porcine or murine or sheep or lambs or pigs or piglets or rabbit or rabbits or cat or cats or dog or dogs or cattle or bovine or monkey or monkeys or trout or marmoset?).ti. and animal experiment/ (1213222)
60. Animal experiment/ not (human experiment/ or human/) (2551334)
61. or/49-60 (4305782)
62. 48 not 61 (5611332)
63. 28 and 62 (3188)
64. limit 63 to english language (3101)
65. 64 not medline.cr. (2547)
66. limit 65 to dc=20220316-20230725 (383)

### PEDro (updated search 2023-08-01

^ For all 2022 update search lines - applied date limit as new records added since 09/11/2020

+ For all 2023 update search lines - applied date limit as new records added since 20/03/2022

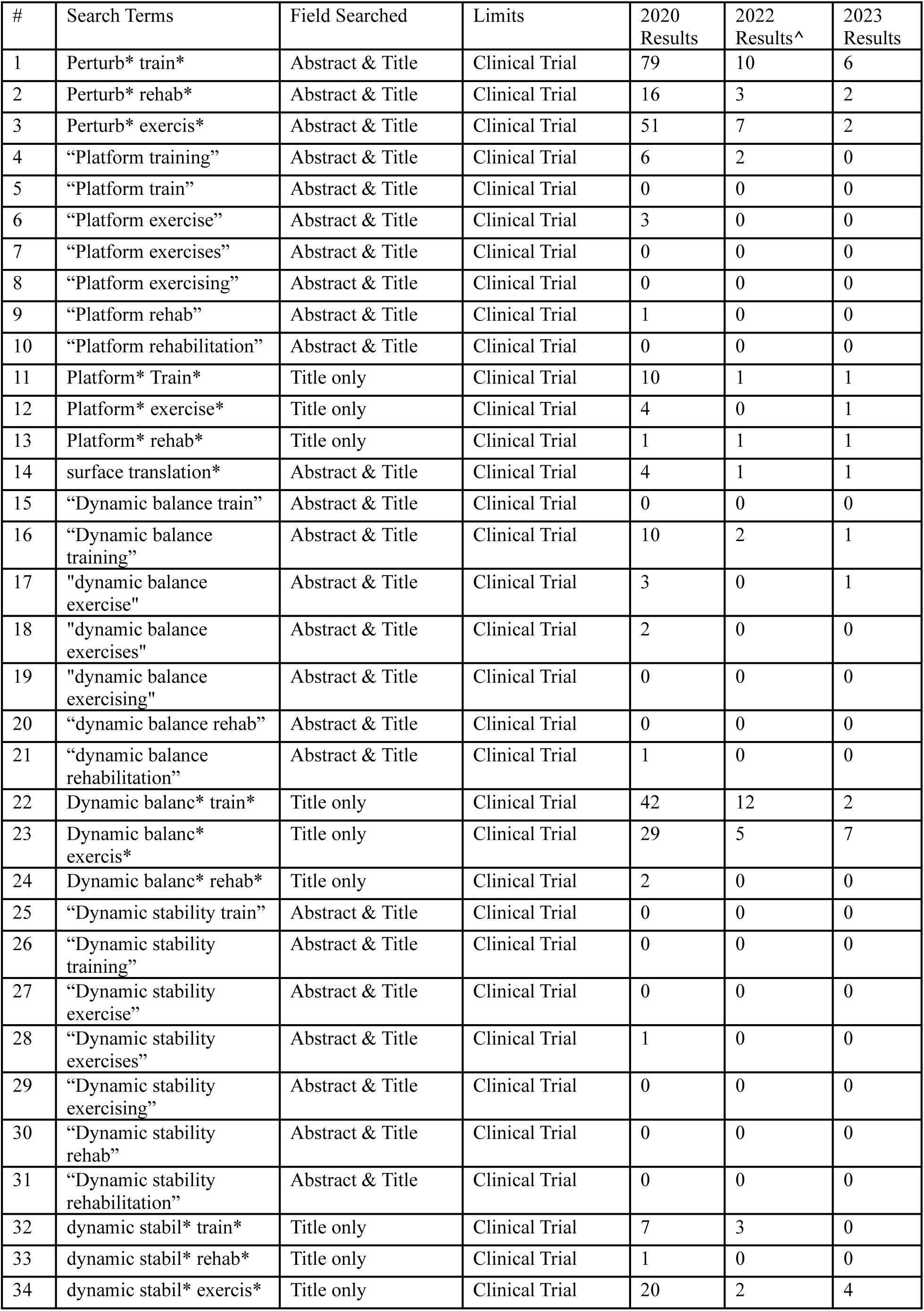
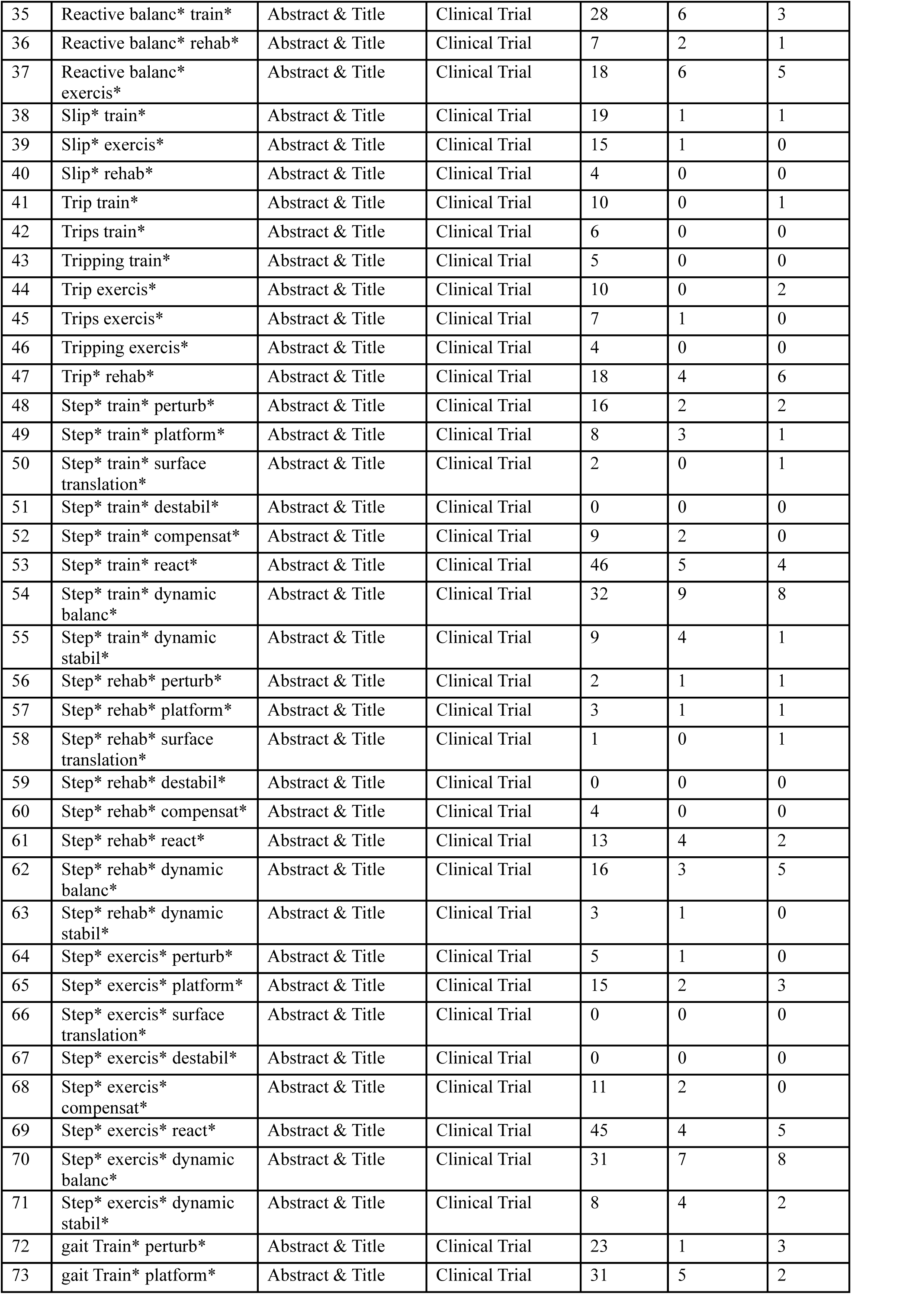
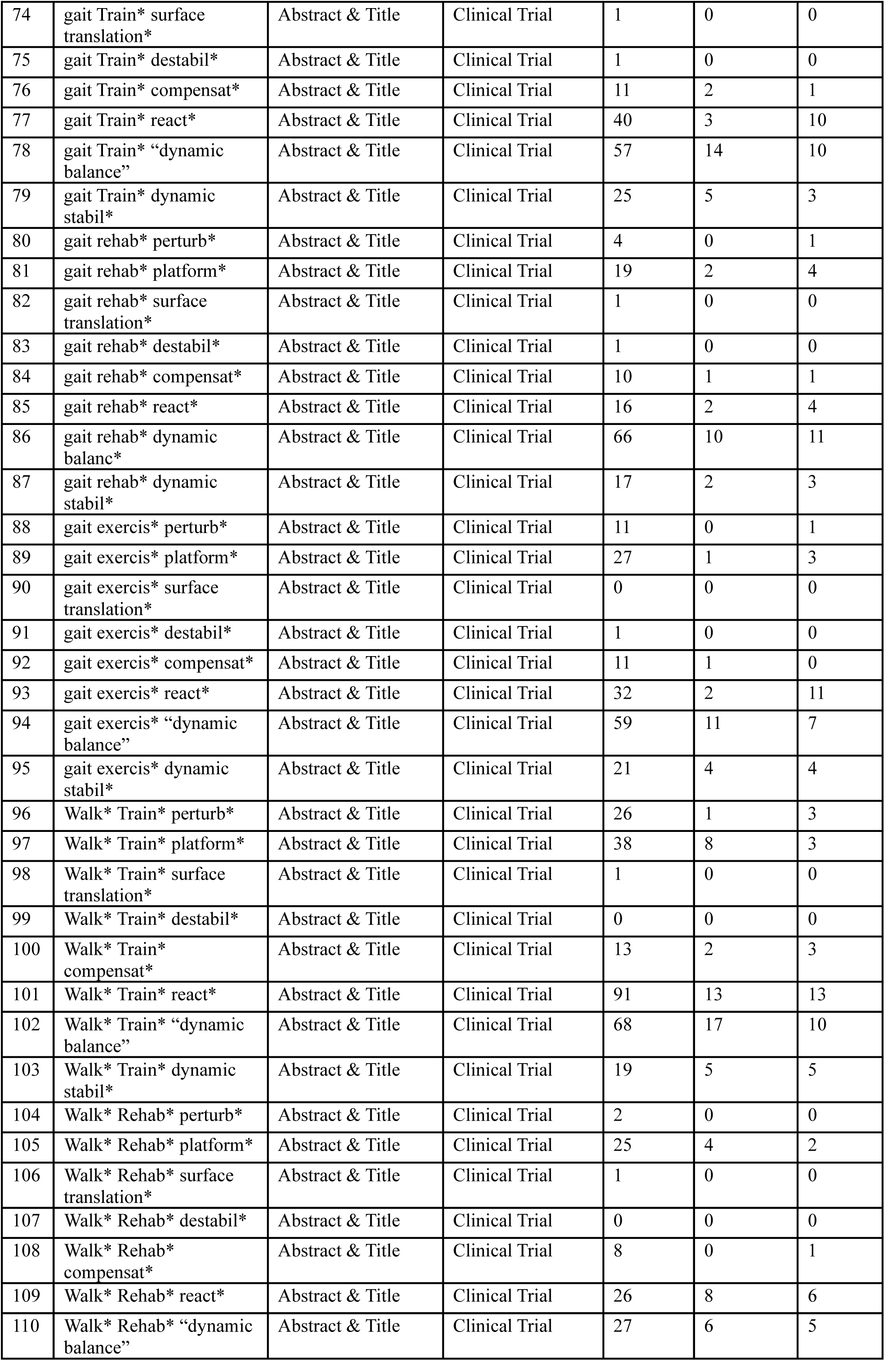
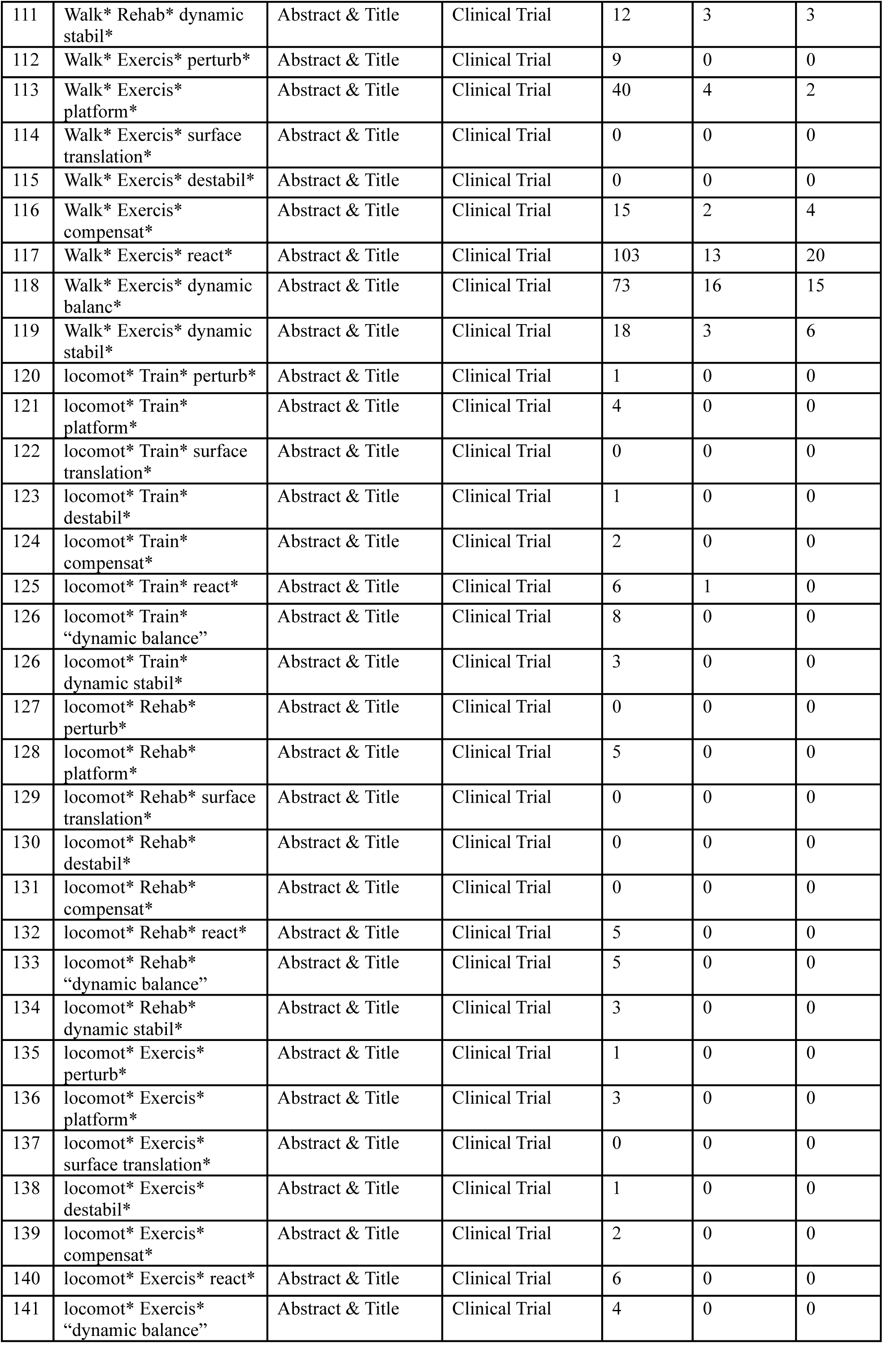
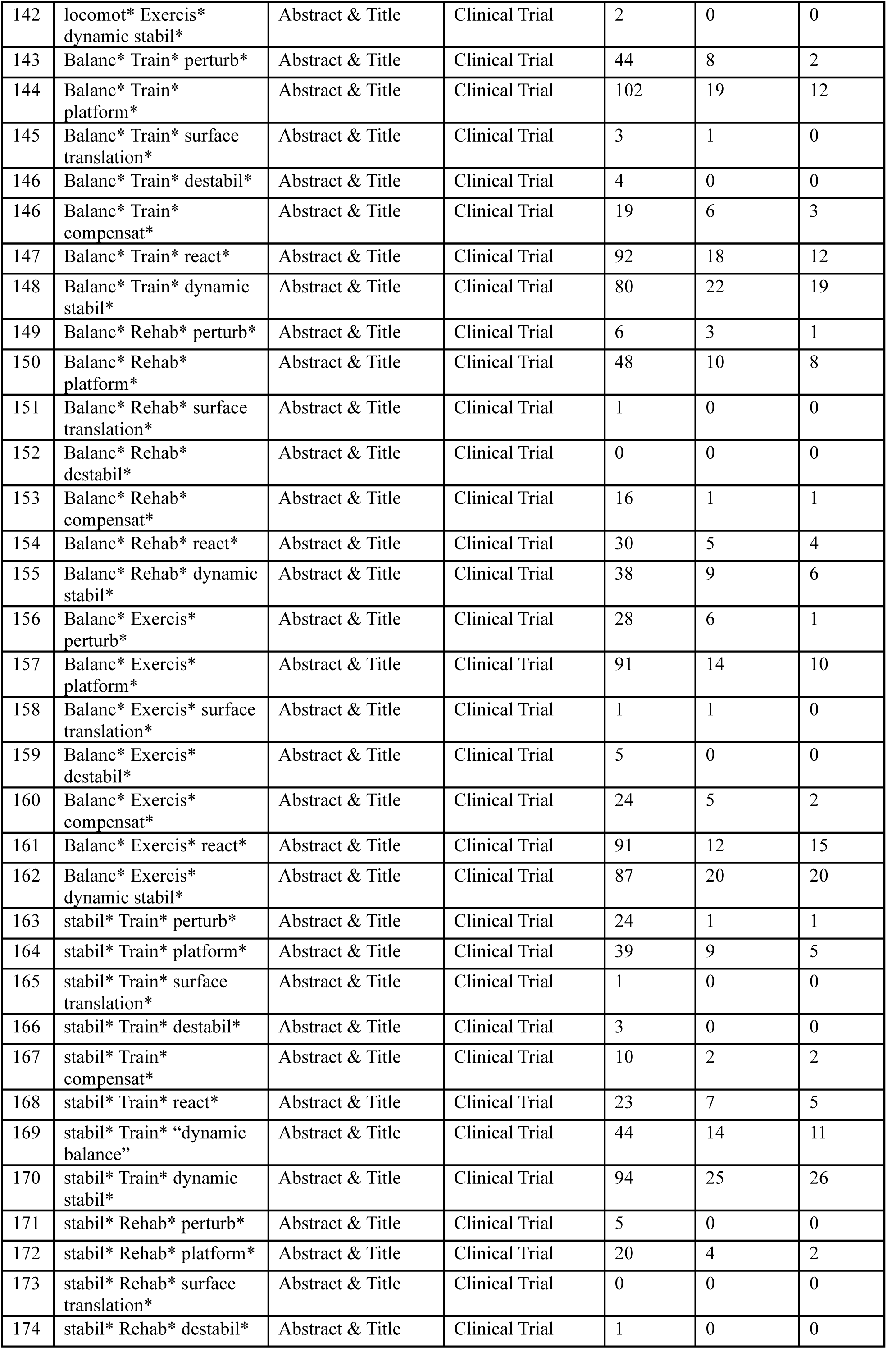
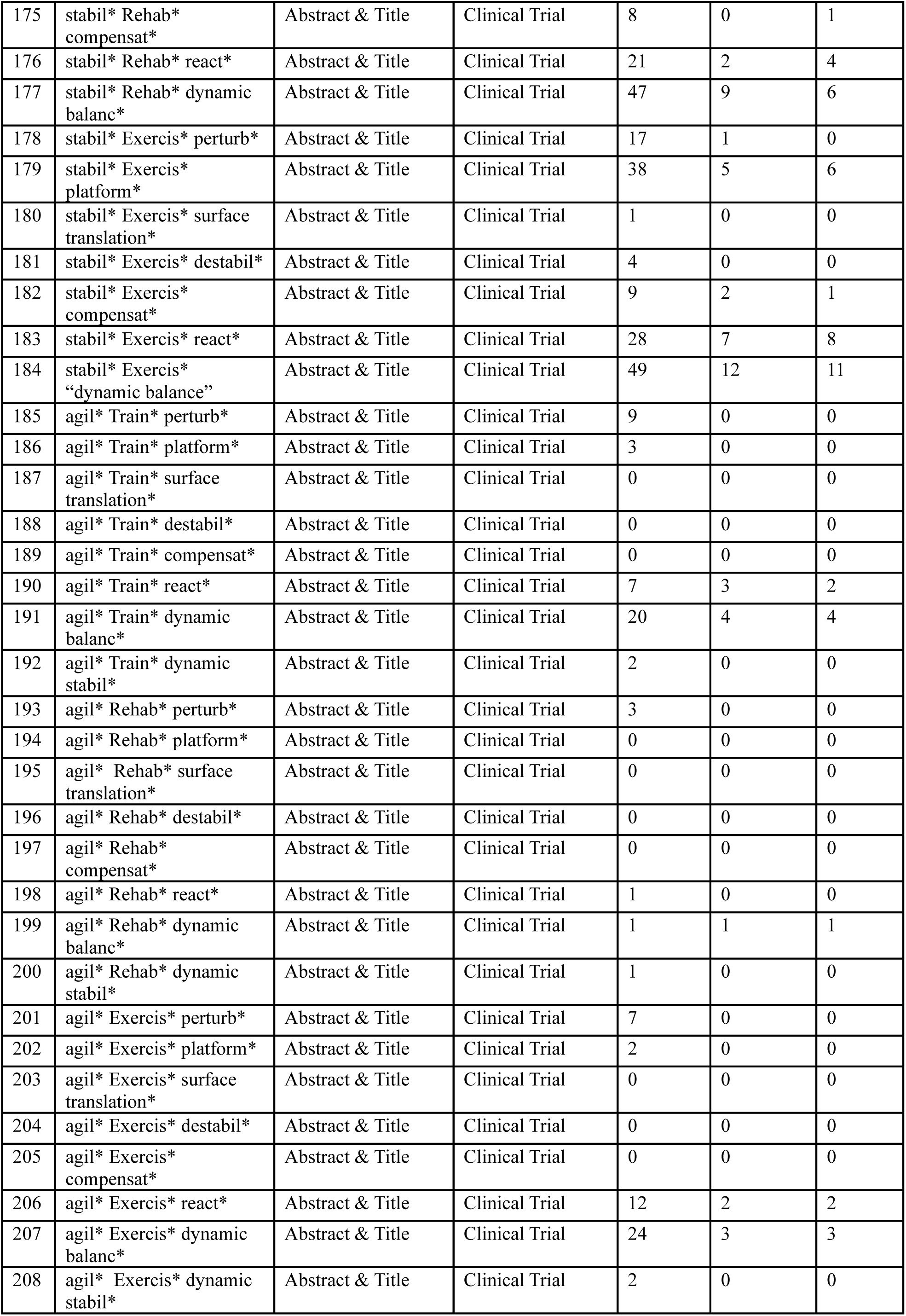

## Appendix 2: Records included in the review

*Primary results papers are indicated with an asterisk

### Shimada 2004

- *Shimada H, Obuchi S, Furuna T, Suzuki T. New intervention program for preventing falls among frail elderly people: the effects of perturbed walking exercise using a bilateral separated treadmill. American Journal of Physical Medicine & Rehabilitation. 2004;83(7):493-499.

### Marigold 2005

- *Marigold DS, Eng JJ, Dawson AS, Inglis JT, Harris JE, Gylfadottir S. Exercise leads to faster postural reflexes, improved balance and mobility, and fewer falls in older persons with chronic stroke. Journal of the American Geriatrics Society. 2005;53(3):416-423.

### Protas 2005

- *Protas EJ, Mitchell K, Williams A, Qureshy H, Caroline K, Lai EC. Gait and step training to reduce falls in Parkinson’s disease. Neurorehabilitation. 2005;20(3):183-190.

### Bieryla 2007

- *Bieryla KA, Madigan Medio-lateral, Nussbaum MA. Practicing recovery from a simulated trip improves recovery kinematics after an actual trip. Gait & Posture. 2007;26(2):208-213.

### Maki 2008

- *Maki BE, Cheng KCC, Mansfield A, et al. Preventing falls in older adults: new interventions to promote more effective change-in-support balance reactions. Journal of Electromyography and Kinesiology. 2008;18(2):243-254.

### Mansfield 2010

- *Mansfield A, Peters AL, Liu BA, Maki BE. Effect of a perturbation-based balance training program on compensatory stepping and grasping reactions in older adults: a randomized controlled trial. Physical Therapy. 2010;90(4):476-491.
- Mansfield A, Peters AL, Liu BA, Maki BE. A perturbation-based balance training program for older adults: study protocol for a randomised controlled trial. BMC Geriatrics. 2007;7:12.
- Mansfield A. Development and evaluation of a perturbation-based balance-training program for older adults [Dissertation/ Thesis]. Toronto, Ontario, Canada: Institute of Medical Science, University of Toronto; 2007.

### Smania 2010

- *Smania N, Corato E, Tinazzi M, et al. Effect of balance training on postural instability in patients with idiopathic Parkinson’s disease. Neurorehabilitation & Neural Repair. 2010;24(9):826-834.

### Parijat 2012

- *Parijat P, Lockhart TE. Effects of moveable platform training in preventing slip-induced falls in older adults. Annals of Biomedical Engineering. 2012;40(5):1111-1121.
- Liu J, Lockhart TE, Parijat P, McIntosh JD, Chiu YP. Comparison of slip training in VR environment and on moveable platform. Biomedical Sciences Instrumentation. 2015;51:189-197.

### Shen 2012

- *Shen X, Mak MKY. Repetitive step training with preparatory signals improves stability limits in patients with Parkinson’s disease. Journal of Rehabilitation Medicine. 2012;44(11):944-949.
- Shen CX, Mak MKY. Effects of 4-week compensatory step training on balance and gait performance in patients with Parkinson’s disease. Movement Disorders. 2010;2):S309.

### Lurie 2013

- *Lurie JD, Zagaria AB, Pidgeon DM, Forman JL, Spratt KF. Pilot comparative effectiveness study of surface perturbation treadmill training to prevent falls in older adults. BMC Geriatrics. 2013;13:49.

### Pai 2014

- *Pai YC, Bhatt T, Yang F, Wang E. Perturbation training can reduce community-dwelling older adults’ annual fall risk: a randomized controlled trial. Journals of Gerontology Series A-Biological Sciences & Medical Sciences. 2014;69(12):1586-1594.

### Schlenstedt 2015

- *Schlenstedt C, Paschen S, Kruse A, Raethjen J, Weisser B, Deuschl G. Resistance versus balance training to improve postural control in parkinson’s disease: a randomized rater blinded controlled study. PLoS ONE. 2015;10(10):e0140584.

### Shen 2015

- *Shen X, Mak MKY. Technology-assisted balance and gait training rreduces falls in patients with Parkinson’s disease: a randomized controlled trial with 12-month follow-up. Neurorehabilitation and neural repair. 2015;29(2):103-111.

### Kurz 2016

- *Kurz I, Gimmon Y, Shapiro A, Debi R, Snir Y, Melzer I. Unexpected perturbations training improves balance control and voluntary stepping times in older adults - a double blind randomized control trial. BMC Geriatrics. 2016;16:58.
- Gimmon Y, Riemer R, Kurz I, Shapiro A, Debbi R, Melzer I. Perturbation exercises during treadmill walking improve pelvic and trunk motion in older adults-A randomized control trial. Archives of Gerontology & Geriatrics. 2018;75:132-138.

### Stieb 2017

- *Steib S, Klamroth S, Gassner H, et al. Perturbation during treadmill training improves dynamic balance and gait in Parkinson’s disease: a single-blind randomized controlled pilot trial. Neurorehabilitation and Neural Repair. 2017;31(8):758-768.
- Pasluosta CF, Steib S, Klamroth S, et al. Acute neuromuscular adaptations in the postural control of patients with Parkinson’s disease after perturbed walking. Frontiers in aging neuroscience. 2017;9:316.
- Klamroth S, Gasner H, Winkler J, et al. Interindividual balance adaptations in response to perturbation treadmill training in persons with Parkinson disease. Journal of Neurologic Physical Therapy. 2019;43(4):224-232
- Steib S, Klamroth S, Gasner H, et al. Exploring gait adaptations to perturbed and conventional treadmill training in Parkinson’s disease: time-course, sustainability, and transfer. Human Movement Science. 2019;64:123-132.
- Klamroth S, Steib S, Gasner H, et al. Immediate effects of perturbation treadmill training on gait and postural control in patients with Parkinson’s disease. Gait & Posture. 2016;50:102-108.
- Gassner H, Steib S, Klamroth S, et al. Perturbation treadmill training improves clinical rating of the motor symptoms gait and postural stability, and sensor-based gait parameters in parkinson’s disease. Gait & posture. 2017;57:345-346.
- Gassner H, Steib S, Klamroth S, et al. Perturbation treadmill training improves clinical characteristics of gait and balance in Parkinson’s disease. Journal of Parkinson’s Disease. 2019;9(2):413-426.

### Mansfield 2018

- *Mansfield A, Aqui A, Danells CJ, et al. Does perturbation-based balance training prevent falls among individuals with chronic stroke? A randomised controlled trial. BMJ Open. 2018;8(8):e021510.
- Schinkel-Ivy A, Huntley AH, Aqui A, Mansfield A. Does perturbation-based balance training improve control of reactive stepping in individuals with chronic stroke? Journal of Stroke & Cerebrovascular Diseases. 2019;28(4):935-943.
- Mansfield A, Aqui A, Centen A, et al. Perturbation training to promote safe independent mobility post-stroke: study protocol for a randomized controlled trial. BMC Neurology. 2015;15:87.

### Aviles 2019

- *Aviles J, Allin LJ, Alexander NB, Van Mullekom J, Nussbaum MA, Madigan Medio-lateral. Comparison of treadmill trip-like training versus tai chi to improve reactive balance among independent older adult residents of senior housing: a pilot controlled trial. Journals of Gerontology Series A-Biological Sciences & Medical Sciences. 2019;74(9):1497-1503.

### Handelzalts 2019

- *Handelzalts S, Kenner-Furman M, Gray G, Soroker N, Shani G, Melzer I. Effects of perturbation-based balance training in subacute persons with stroke: a randomized controlled trial. Neurorehabilitation & Neural Repair. 2019;33(3):213-224.

### Okubo 2019

- *Okubo Y, Sturnieks DL, Brodie MA, Duran L, Lord SR. Effect of reactive balance training involving repeated slips and trips on balance recovery among older adults: a blinded randomized controlled trial. Journals of Gerontology Series A-Biological Sciences & Medical Sciences. 2019;74(9):1489-1496.

### Esmaeili 2020

- *Esmaeili V, Juneau A, Dyer JO, et al. Intense and unpredictable perturbations during gait training improve dynamic balance abilities in chronic hemiparetic individuals: a randomized controlled pilot trial. Journal of Neuroengineering & Rehabilitation. 2020;17(1):79.

### Lurie 2020

- *Lurie JD, Zagaria AB, Ellis L, et al. Surface perturbation training to prevent falls in older adults: a highly pragmatic, randomized controlled trial. Physical Therapy. 2020;100(7):1153-1162.

### Rieger 2020

- *Rieger MM, Papegaaij S, Pijnappels M, Steenbrink F, van Dieen JH. Transfer and retention effects of gait training with anterior-posterior perturbations to postural responses after medio-lateral gait perturbations in older adults. Clinical Biomechanics. 2020;75:104988.

### Rogers 2021

- *Rogers MW, Creath RA, Gray V, et al. Comparison of lateral perturbation-induced step training and hip muscle strengthening exercise on balance and falls in community-dwelling older adults: a blinded randomized controlled trial. Journals of Gerontology Series A-Biological Sciences & Medical Sciences. 08 13 2021;76(9):e194-e202.

### Unger 2021

- *Unger J, Chan K, Lee JW, et al. The effect of perturbation-based balance training and conventional intensive balance training on reactive stepping ability in individuals with incomplete spinal cord injury or disease: a randomized clinical trial. Front Neurol. 2021;12:620367.
- Unger J, Chan K, Scovil CY, et al. Intensive balance training for adults with incomplete spinal cord injuries: protocol for an assessor-blinded randomized clinical trial. Physical Therapy. 2019;99(4):420-427.

### Monjezi 2022

- *Monjezi S, Molhemi F, Shaterzadeh-Yazdi M-J, et al. Perturbation-based balance training to improve postural responses and falls in people with multiple sclerosis: a randomized controlled trial. Disability and rehabilitation. 2022(9207179, a8i):1-7.

### Wang 2022

- *Wang Y, Wang S, Liu X, Lee A, Pai YC, Bhatt T. Can a single session of treadmill-based slip training reduce daily life falls in community-dwelling older adults? A randomized controlled trial. Aging Clinical & Experimental Research. 2022;02:02.
- Wang Y, Bhatt T, Liu X, et al. Can treadmill-slip perturbation training reduce immediate risk of over-ground-slip induced fall among community-dwelling older adults? Journal of Biomechanics. 2019;84:58-66.

### Brull 2023

- *Brull L, Hezel N, Arampatzis A, Schwenk M. Comparing the Effects of Two Perturbation-Based Balance Training Paradigms in Fall-Prone Older Adults: A Randomized Controlled Trial. Gerontology. 2023;69(7):910-922.

### de Souza 2023

- *de Souza CR, Avila de Oliveira J, Takazono PS, et al. Perturbation-based balance training leads to improved reactive postural responses in individuals with Parkinson’s disease and freezing of gait. The European journal of neuroscience. 2023;57(12):2174-2186.
- Effect of training with instability on a platform in patients with Parkinson’s disease. Effect of postural perturbation training on the ability to recover body balance in individuals with Parkinson’s disease with freezing of gait 2018; http://ovidsp.ovid.com/ovidweb.cgi?T=JS&PAGE=reference&D=cctr&NEWS=N&AN=CN-02433746.

### Gerards 2023

- *Gerards M, Marcellis R, Senden R, et al. The effect of perturbation-based balance training on balance control and fear of falling in older adults: a single-blind randomised controlled trial. BMC Geriatrics. 2023;23(1):305.
- Gerards MHG, Marcellis RGJ, Poeze M, Lenssen AF, Meijer K, de Bie RA. Perturbation-based balance training to improve balance control and reduce falls in older adults - study protocol for a randomized controlled trial. BMC Geriatrics. 2021;21(1):9.

### Nørgaard 2023

- *Norgaard JE, Andersen S, Ryg J, et al. Effect of treadmill perturbation-based balance training on fall rates in community-dwelling older adults: a randomized clinical trial. JAMA Netw Open. 2023;6(4):e238422.
- Norgaard JE, Andersen S, Ryg J, et al. Effects of treadmill slip and trip perturbation-based balance training on falls in community-dwelling older adults (STABILITY): study protocol for a randomised controlled trial. BMJ Open. 2022;12(2):e052492.

### Okubo 2023

- *Okubo Y, Mohamed Suhaimy MSB, Hoang P, et al. Training reactive balance using trips and slips in people with multiple sclerosis: a blinded randomised controlled trial. Multiple Sclerosis and Related Disorders. 2023;73(101580247):104607.
- MS-SAFE: stepping to avoid fall events in multiple sclerosis. Training protective stepping responses to trips and slips in people with multiple sclerosis 2018; http://ovidsp.ovid.com/ovidweb.cgi?T=JS&PAGE=reference&D=cctr&NEWS=N&AN=CN-02445220.

### Rieger 2024

- *Rieger MM, Papegaaij S, Steenbrink F, van Dieën JH, Pijnappels M. Effects of perturbation-based treadmill training on balance performance, daily-life gait and falls in older adults; REACT randomized controlled trial. Phys Ther. 2024;104(1):pzad136
- Rieger MM, Papegaaij S, Steenbrink F, van Dieen JH, Pijnappels M. Perturbation-based gait training to improve daily life gait stability in older adults at risk of falling: protocol for the REACT randomized controlled trial. BMC Geriatrics. 2020;20(1):167.
- Perturbation-based gait training. Perturbation-based gait training: evaluation of a tool to improve balance and gait in older people at risk of falling 2019; http://ovidsp.ovid.com/ovidweb.cgi?T=JS&PAGE=reference&D=cctr&NEWS=N&AN=CN-02433791.

## Appendix 3: Excluded studies that appear to meet eligibility criteria

### No falls or reactive balance outcome

- Toole T, Hirsch MA, Forkink A, Lehman DA, Maitland CG. The effects of a balance and strength training program on equilibrium in Parkinsonism: A preliminary study. Neurorehabilitation. 2000;14(3):165-74.
- Allin LJ, Brolinson PG, Beach BM, Kim S, Nussbaum MA, Roberto KA, et al. Perturbation-based balance training targeting both slip- and trip-induced falls among older adults: a randomized controlled trial. BMC Geriatrics. 2020;20(1):205.
- Rogge AK, Hotting K, Nagel V, Zech A, Holig C, Roder B. Improved balance performance accompanied by structural plasticity in blind adults after training. Neuropsychologia. 2019;129:318-30.
- Bonni S, Ponzo V, Tramontano M, Martino Cinnera A, Caltagirone C, Koch G, et al. Neurophysiological and clinical effects of blindfolded balance training (BBT) in Parkinson’s disease patients: a preliminary study. European journal of physical & rehabilitation medicine. 2019;55(2):176-82.
- Wrisley DM, Stephens MJ. The effects of rotational platform training on balance and ADLs. Annual International Conference Of The IEEE Engineering In Medicine And Biology Society. 2011;2011:3529-32.
- Melzer I, Oddsson L. Improving balance control and self-reported lower extremity function in community-dwelling older adults: a randomized control trial. Clinical Rehabilitation. 2013;27(3):195-206.
- Halvarsson A, Oddsson L, Olsson E, Faren E, Pettersson A, Stahle A. Effects of new, individually adjusted, progressive balance group training for elderly people with fear of falling and tend to fall: a randomized controlled trial. Clinical Rehabilitation. 2011;25(11):1021-31.
- Rogers MW, Johnson ME, Martinez KM, Mille Medio-lateral, Hedman LD. Step training improves the speed of voluntary step initiation in aging. Journals of Gerontology Series A-Biological Sciences & Medical Sciences. 2003;58(1):46-51.
- Gandolfi M, Geroin C, Dimitrova E, Boldrini P, Waldner A, Bonadiman S, et al. Virtual reality telerehabilitation for postural instability in Parkinson’s disease: a multicenter, single-blind, randomized, controlled trial. BioMed Research International. 2017(7962826).
- Beling J, Roller M. Multifactorial intervention with balance training as a core component among fall-prone older adults. Journal of Geraitric Physical Therapy. 2009;32(3):125-33.
- Halvarsson A, Franzén E, Farén E, Olsson E, Oddsson L, Ståhle A. Long-term effects of new progressive group balance traning for elderly people with increased risk of falling - a randomized controlled trial. Clin Rehabil. 2013;27(5):450-8.
- Batcir S, Livne Y, Lev Lehman R, Edelman S, Schiller L, Lubovsky O, et al. Development and piloting of a perturbation stationary bicycle robotic system that provides unexpected lateral perturbations during bicycling (the PerStBiRo system). BMC Geriatrics. 2021;21(1):71.
- Batcir S, Lubovsky O, Bachner YG, Melzer I. The effects of bicycle simulator training on anticipatory and compensatory postural control in older adults: study protocol for a single-blind randomized controlled trial. Frontiers in neurology. 2020;11:614664.
- Bhatt T, Wang Y, Wang S, Kannan L. Perturbation training for fall-risk reduction in healthy older adults: interference and generalization to opposing novel perturbations post intervention. Front. 2021;3:697169.
- Kumar C, Pathan N. Effectiveness of manual perturbation exercises in improving balance, function and mobility in stroke patients: a randomized controlled trial. Journal of Novel Physiotherapies 2016;6(2):284. 2016.
- Adeniyi A, Stramel DM, Rahman D, Rahman M, Yadav A, Zhou J, et al. Utilizing mobile robotics for pelvic perturbations to improve balance and cognitive performance in older adults: a randomized controlled trial. Research square. 2023(101768035).
- Pereira DB, Souza TSd, Fuzinato CT, Hagihara RJ, Ribeiro Antero-posterior. Effect of a programme of muscular endurance, balance and gait exercises with and without the use of flexible and minimalist shoes in older women with medial knee osteoarthritis: study protocol for a randomised controlled trial. BMJ Open. 2022;12(9):e061267.
- Diniz-Sousa F, Granja T, Boppre G, Veras L, Devezas V, Santos-Sousa H, et al. Effects of a multicomponent exercise training program on balance following bariatric surgery. International Journal of Sports Medicine. 2022;43(9):818-24.
- PBBT and WBV effect on balance and gait in stroke. 2022; https://clinicaltrials.gov/study/NCT05588661
- Diniz-Sousa F, Veras L, Boppre G, Sa-Couto P, Devezas V, Santos-Sousa H, et al. The effect of an exercise program on bone health after bariatric surgery: a randomized controlled trial. Journal of Bone and Mineral Research. 2021;36(3):489-99.

### Study still ongoing

- Barzideh A, Marzolini S, Danells C, Jagroop D, Huntley AH, Inness EL, et al. Effect of reactive balance training on physical fitness poststroke: study protocol for a randomised non-inferiority trial. BMJ Open. 2020;10(6):e035740.
- Miller KJ, Hunt MA, Pollock CL, Bryant D, Garland SJ. Protocol for a randomized controlled clinical trial investigating the effectiveness of Fast muscle Activation and Stepping Training (FAST) for improving balance and mobility in sub-acute stroke. BMC Neurology. 2014;14:187.
- Trampisch US, Petrovic A, Daubert D, Wirth R. Fall prevention by reactive balance training on a perturbation treadmill: is it feasible for prefrail and frail geriatric patients? A pilot study. European geriatric medicine. 2023(101533694).
- Domingos J, Dean J, Fernandes JB, Ramos C, Grunho M, Proenca L, et al. Lisbon Intensive Falls Trampoline Training (LIFTT) program for people with Parkinson’s for balance, gait, and falls: study protocol for a randomized controlled trial. Trials. 2023;24(1):101.
- PNF vs pertubation based balance training in subacute stroke patients. 2023; https://clinicaltrials.gov/study/NCT05856045
- Retraining of balance, nerve glide exercises, resistance training, and instruction in daily living skills are effective treatments for chemotherapy-induced peripheral neuropathy in cancer survivor. 2023; https://ctri.nic.in/Clinicaltrials/pmaindet2.php?EncHid=NzY4MTU=&Enc=&userName=CT <u>RI/2023/03/050356</u>
- Treadmill perturbation training for fall prevention after total knee replacement. 2023; https://clinicaltrials.gov/ct2/show/NCT05736666
- The effect of exercise therapy on balance in patients with multiple sclerosis. 2023; http://ovidsp.ovid.com/ovidweb.cgi?T=JS&PAGE=reference&D=cctr&NEWS=N&AN=CN-02521031
- High-intensity, dynamic-stability gait training in people with multiple sclerosis. 2023; https://clinicaltrials.gov/study/NCT05735691
- Perturbation-Based Balance Training for stroke patient. 2022; https://trialsearch.who.int/Trial2.aspx?TrialID=CTRI/2022/11/047124
- ReacStep Study: step Training program for improving fall risk and cognition in older adults. Effects of a 6-week step training program on fall risk and cognition in older adults: a blinded randomised controlled trial. 2022; https://www.anzctr.org.au/Trial/Registration/TrialReview.aspx?id=382207
- Is running re-education group more effective than high level balance group in reducing falls in the elderly living in the community? 2013; https://www.anzctr.org.au/Trial/Registration/TrialReview.aspx?id=365359

### Unable to obtain data or study details

- Konig M, Epro G, Seeley J, Potthast W, Karamanidis K. Retention and generalizability of balance recovery response adaptations from trip perturbations across the adult life span. Journal of Neurophysiology. 2019;122(5):1884-93. Note: Data for the reactive balance control measure are reported in a figure in the manuscript. We were unable to contact the corresponding author to obtain means and standard deviations for data reported in the figure; contact information were invalid, and we were unable to find an alternative contact.
- Manko G, Pieniazek M, Tim S, Jekielek M. The effect of Frankel’s stabilization exercises and stabilometric platform in the balance in elderly patients: a randomized clinical trial. Medicina. 2019;55(9):11. Note: Full details of the intervention were not reported in the manuscript. The Performance Oriented Mobility Assessment was completed as a measure of reactive balance control, but the data were not reported in way that could be incorporated into the meta-analysis. The corresponding author was contacted, but we did not receive a response.
- Bierbaum S, Peper A, Arampatzis A. Exercise of mechanisms of dynamic stability improves the stability state after an unexpected gait perturbation in elderly. Age. 2013;35(5):1905-15. Note: Data for the reactive balance control measure are reported in a figure in the manuscript. The corresponding author was contacted to obtain the values for data reported in the figure, but we did not receive a response.
- Morgan P, Murphy A, Opheim A. The safety and feasibility of an intervention to improve balance dysfunction in ambulant adults with cerebral palsy: a pilot randomized controlled trial. Clin Rehabil. 2015;29(9):907-19. Note: Full intervention details were not reported in the manuscript. The corresponding author was unable to provide these details as study records had been destroyed per institutional policies.
- Arampatzis A, Peper A, Bierbaum S. Exercise of mechanisms for dynamic stability control increases stability performance in the elderly. J Biomech. 2011;44(1):52-8. Note: Data for the reactive balance control measure are reported in a figure in the manuscript. The corresponding author was contacted to obtain the values for data reported in the figure, but we did not receive a response.
- Improving neural function through targeted exercise in older adults. 2017; https://uat.anzctr.org.au/Trial/Registration/TrialReview.aspx?id=373862 Note: This is a trial registry record. The trial was completed in June 2014 according to data in the registry. We were unable to find a published study reporting the results. Contact information in the registry was invalid, and we were unable to find an alternative contact.

### Not in English

- Obuchi S, Kojima M, Shiba Y, Shimada H, Suzuki T. (A randomized controlled trial of a treadmill training with the perturbation to improve the balance performance in the community dwelling elderly subjects) [Japanese]. Nippon Ronen Igakkai Zasshi [Japanese Journal of Geriatrics]. 2004;41(3):321-7.
- Lin C, Chen A, Jiang Y. Effect of virtual reality balance game training on balance function in patients with Parkinson’s disease. Theory Pract Rehab China. 2016;22:1059-63.

### Not all RBT participants received RBT

- de Rooij IJM, van de Port IGL, Punt M, Abbink-van Moorsel PJM, Kortsmit M, van Eijk RPA, et al. Effect of virtual reality gait training on participation in survivors of subacute stroke: a randomized controlled trial. Physical Therapy. 2021;101(5):04. Note: The corresponding author was contacted regarding details of the intervention. While the virtual reality intervention may have included some reactive balance training, not all participants in this group received reactive balance training. The authors did not document details of training received.

